# Relationships Between Brain Functional Connectivity and Resting Cardiac Autonomic Profiles in Functional Neurological Disorder: A Pilot Study

**DOI:** 10.64898/2026.01.06.26343455

**Authors:** Cristina Bleier, Andrew J. Guthrie, Jessica Ranford, Julie MacLean, Ellen Godena, Julie Maggio, Sara A. Finkelstein, Ibai Diez, Christiana Westlin, Karen S. Quigley, David L. Perez

## Abstract

**Background:** Functional neurological disorder (FND) is associated with alterations in functional brain networks, yet relationships between peripheral autonomic physiology and brain architecture remain poorly characterized. This pilot study examined associations between cardiac autonomic metrics and resting-state functional connectivity (rsFC) in FND.

**Methods:** Twenty females with FND and 23 age-matched female psychiatric controls (PCs) completed questionnaires, 10-min resting photoplethysmography recordings, and same-day resting-state fMRI. Interbeat interval (IBI) and heart rate variability (HRV) metrics were extracted. Whole-brain rsFC was quantified using weighted-degree [centrality]. Within-group analyses tested associations between cardiac autonomic metrics and weighted-degree rsFC separately in FND and PC cohorts, adjusting for age, head motion, and antidepressant/β-blocker use – while applying a cluster-wise correction.

**Results:** Cardiac (IBI and HRV) metrics did not differ between FND and PC cohorts, and these metrics did not correlate with FND symptom severity, somatic symptom burden, affective symptoms, or childhood trauma. In FND, shorter IBI (i.e., faster resting heart rate) correlated with increased weighted-degree rsFC in bilateral supplementary motor area (SMA) and right precentral/superior frontal regions, whereas higher HRV primarily correlated with decreased weighted-degree rsFC in the bilateral SMA, mid-cingulate cortex, and right amygdala, anterior insula, and lateral orbitofrontal cortex. In PCs, autonomic–rsFC associations were more spatially restricted to the anterior/mid-cingulate and SMA.

**Conclusion:** In FND, individual differences in resting autonomic physiology related to the centrality of brain areas that are part of the central autonomic, salience, and allostatic-interoceptive networks. These findings suggest that the relationship between autonomic physiology and network architecture may be important in FND.

## INTRODUCTION

Functional Neurological Disorder (FND) is a prevalent, costly, and potentially disabling condition at the intersection of neurology and psychiatry (Aybek and Perez, 2022; Hallett et al., 2022; O’Mahony et al., 2023). Epidemiologic studies demonstrate that FND represents a substantial proportion of outpatient neurology referrals, and is associated with $2 billion annually in emergency department and acute inpatient hospital expenditures in the United States (Finkelstein et al., 2025; Stephen et al., 2025). Clinically, patients with FND exhibit heterogeneous combinations of motor, sensory, and cognitive symptoms that contribute to reductions in health-related quality of life (Anderson et al., 2007; Jones et al., 2016). Concurrent mood, anxiety, and trauma-related symptoms are also common in this population (Butler et al., 2021; Gray et al., 2020). Importantly, clinical practice has shifted from FND as a diagnosis of exclusion to identifying positive “rule-in’’ examination signs (e.g., tremor entrainment, Hoover’s sign), allowing for expedited diagnosis and earlier treatment (LaFrance et al., 2013; Perez et al., 2021a). These advances have catalyzed interest in clarifying the neural circuits and other physiological mechanisms implicated in the pathophysiology of FND.

Resting-state functional MRI (rs-fMRI) studies in FND have identified altered intrinsic brain organization across multiple distributed large-scale brain networks (Baizabal-Carvallo et al., 2019; Homayoun et al., 2025; Perez et al., 2021b; Yeo et al., 2011). Several investigations converge on alterations in the salience, somatomotor, default mode, and frontoparietal networks (Li et al., 2015a; van der Kruijs et al., 2012; Weber et al., 2022; Wegrzyk et al., 2018; Westlin et al., 2025a). In particular, FND cohort studies identify stronger coupling between regions of the salience/ventral attention network—especially the anterior insula, dorsal anterior cingulate cortex, and the amygdala—and motor control areas (Demartini et al., 2021; Diez et al., 2019; Li et al., 2015a, 2015b; Morris et al., 2017; Seeley, 2019; van der Kruijs et al., 2012). Whole-brain connectivity analyses further show increased crosstalk [i.e., *between-network* connectivity] among salience, somatomotor, and default mode networks, the magnitude of which correlated with individual differences in functional somatic symptom severity (Westlin et al., 2025a). Relatedly, these altered connectivity patterns in FND implicate multimodal integration hubs—including the anterior insula, anterior and mid-cingulate cortices, and temporoparietal junction—brain areas involved in integrating interoceptive, affective, and cognitive signals with motor control ones (Mavroudis et al., 2024; Sepulcre et al., 2012). Many of these salience network and multimodal integration hubs—especially the anterior insula, anterior/mid-cingulate cortex, and amygdala—are also components of the central autonomic network (CAN) (Benarroch, 1993; Cechetto and Shoemaker, 2009; Quadt et al., 2022; Saper, 2002) and the allostatic-interoceptive network (Kleckner et al., 2017; Zhang et al., 2025). Within the CAN and allostatic-interoceptive network, distributed cortical, limbic, midbrain and other brainstem regions coordinate autonomic regulation. This anatomical and functional overlap suggests that contextualizing relationships between resting-state functional connectivity (rsFC) and cardiac autonomic metrics may advance our neurobiological understanding of FND.

In FND, characterization of cardiac autonomic profiles is also an area of active research. Individuals with paroxysmal FND events frequently endorse cardiac autonomic symptoms (e.g., racing heart rate [HR]) as a bodily ‘warning sign’ prior to onset of an episode (Goetz et al., 2013; Goldstein and Mellers, 2006; Indranada et al., 2019). This phenomena has been termed “panic attack without panic” – characterized by individuals experiencing some of the physical symptoms associated with panic attacks without a panic-related emotional experience (Goldstein and Mellers, 2006; Jungilligens et al., 2022). Although findings across cohorts are heterogeneous (particularly in adults), several observations point towards altered cardiac autonomic profiles in FND relative to controls (Bakvis et al., 2009; Demartini et al., 2016; Kozlowska et al., 2015, 2017, 2018; Maurer et al., 2016; Ponnusamy et al., 2011; Radmanesh et al., 2020; Sawchuk et al., 2020). A 2022 meta-analysis by Paredes-Echeverri et al. (2022) found higher resting HRs in patients with FND across pediatric and adult cohorts compared to healthy controls (HCs), along with a tendency towards reduced heart rate variability (HRV) at rest (Paredes-Echeverri et al., 2022). Autonomic metrics in some studies have also been shown to co-vary with clinically meaningful variables — including symptom severity, subjective distress, anxiety, and illness duration — further underscoring the importance of autonomic profiles in FND (Allendorfer et al., 2019; Kozlowska et al., 2015). In the pediatric FND literature, several brain imaging studies have probed the neural correlates of cardiac autonomic profiles – principally measured through resting HR. For example, differences in somatosensory cortex-to-cerebellar network connectivity were related to resting HR values in pediatric FND (Rai et al., 2022). RsFC profiles across insular sub-regions also exhibited trend-level correlations with resting HR (Walpola et al., 2025). To our knowledge, no studies in adults with FND have investigated relationships between resting cardiac autonomic metrics and resting-state functional brain organization.

In this pilot study, we examined out-of-scanner cardiac autonomic metrics at rest in females with FND compared to age and sex-matched psychiatric controls (PCs). We focused on peripheral autonomic measurements, including IBI (inverse of HR) and HRV. We subsequently investigated how individual differences in IBI and HRV related to whole-brain, weighted-degree [global centrality] derived from rs-fMRI – with analyses performed separately in FND and PC cohorts. Given that the CAN and allostatic-interoceptive networks include brain areas implicated in the pathophysiology of FND, we hypothesized that shorter resting IBI (i.e., higher resting HR) and lower resting HRV would relate to cingulo-insular and amygdala rsFC profiles. We also hypothesized that a subset of these functional connectivity-autonomic relationships would be distinct in patients with FND compared to PCs, in keeping with partially overlapping and distinct neurobiological mechanisms in these two populations (Diez et al., 2021; Westlin et al., 2025a). Given that medications can influence autonomic physiology, a secondary aim was to explore the impact of antidepressants, beta-blockers, α-adrenergic blockers, and stimulant medications on within-group associations between rsFC and cardiac autonomic profiles.

## METHODS

### Participants

Twenty female participants with FND [mean age (SD) = 35.9±13.2 years; **Table 1**] were prospectively recruited from the Massachusetts General Hospital FND Unit between October 2022 and March 2025. Rule-in diagnoses were established based on positive signs, semiological features (e.g., tight eye closure at event onset, asynchronous limb shaking movements), and electroencephalogram data, when applicable (i.e., for functional seizures only) (LaFrance et al., 2013). The FND cohort was ‘mixed’, meaning individuals had a range of phenotypes; a transdiagnostic approach was taken given that many individuals with FND exhibit mixed symptoms cross-sectionally and/or develop distinct FND symptoms longitudinally (McKenzie et al., 2011; Tinazzi et al., 2020). The FND cohort included 15 individuals with functional motor disorder and seven people with functional seizures; two participants met diagnostic criteria for both phenotypes (see **Supplementary Table 1** for a complete description of participant phenotypes). Exclusion criteria included major neurological comorbidities (e.g., epilepsy, Parkinson’s disease), known brain MRI abnormalities, poorly controlled medical problems with known central nervous system consequences, active illicit substance use disorder, history of psychosis, and/or active suicidality. One additional female with FND was enrolled but excluded following outlier inspection of mean weighted-degree values (defined as having a mean weighted-degree at the whole-brain level exceeding more than 1.5 times the upper or lower interquartile range values). Males with FND were also excluded from this pilot, given the limited number of such individuals who have participated in dual brain MRI and autonomic data collection to date (n=3). This autonomic-focused pilot study is part of a larger project from which cross-sectional structural MRI and rs-fMRI data have been previously published (Westlin et al., 2025a, 2025b, 2025c); no autonomic data for this cohort has been previously published.

**Table 1.**
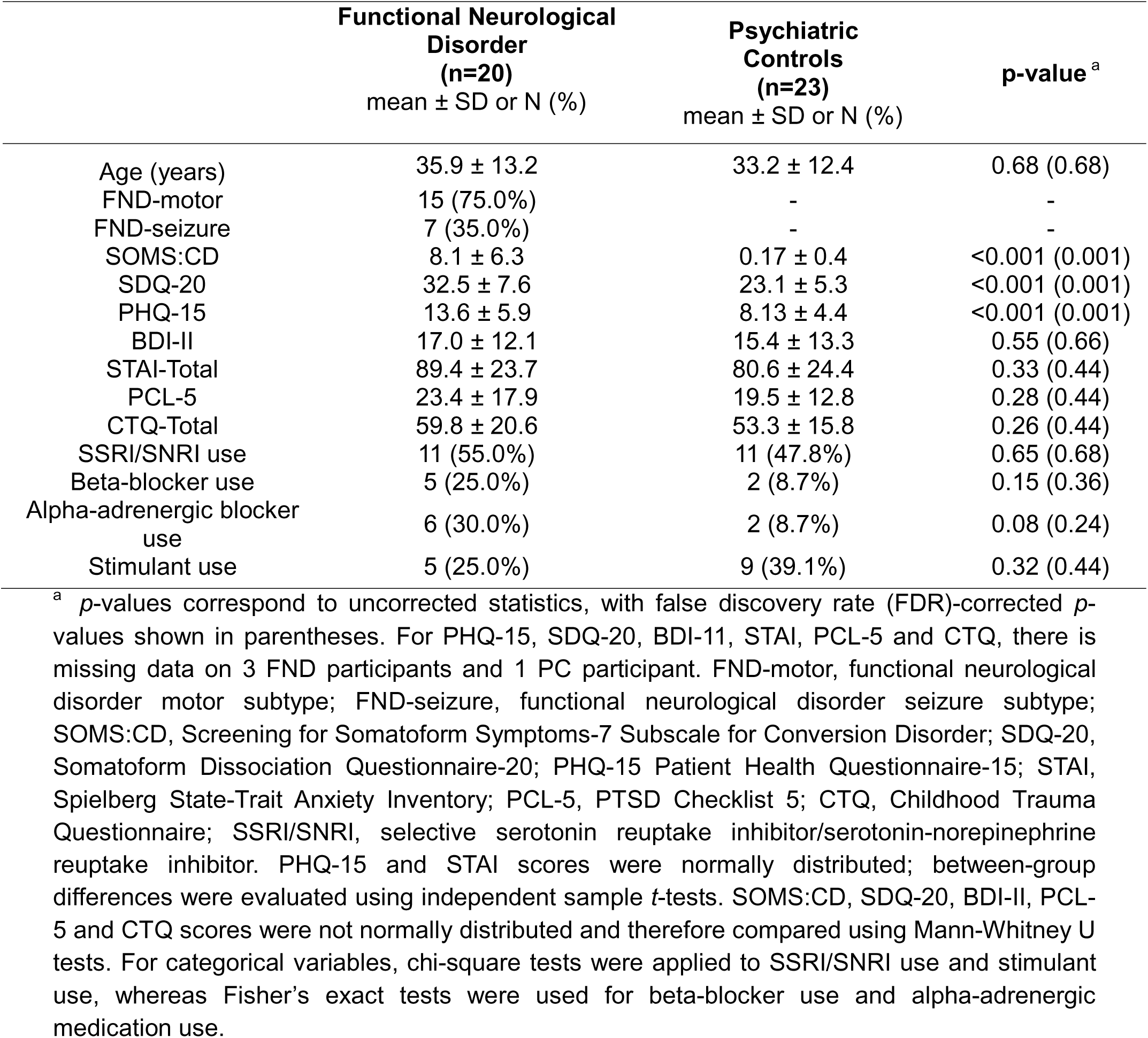
Clinical characteristics of females with functional neurological disorder (FND) versus psychiatric control (PC) cohort.

| | **Functional Neurological Disorder**<br>**(n=20)**<br>mean ± SD or N (%) | **Psychiatric Controls**<br>**(n=23)**<br>mean ± SD or N (%) | **p-value<sup>a</sup>** |
| --- | --- | --- | --- |
| Age (years) | 35.9 ± 13.2 | 33.2 ± 12.4 | 0.68 (0.68) |
| FND-motor | 15 (75.0%) | - | - |
| FND-seizure | 7 (35.0%) | - | - |
| SOMS:CD | 8.1 ± 6.3 | 0.17 ± 0.4 | <0.001 (0.001) |
| SDQ-20 | 32.5 ± 7.6 | 23.1 ± 5.3 | <0.001 (0.001) |
| PHQ-15 | 13.6 ± 5.9 | 8.13 ± 4.4 | <0.001 (0.001) |
| BDI-II | 17.0 ± 12.1 | 15.4 ± 13.3 | 0.55 (0.66) |
| STAI-Total | 89.4 ± 23.7 | 80.6 ± 24.4 | 0.33 (0.44) |
| PCL-5 | 23.4 ± 17.9 | 19.5 ± 12.8 | 0.28 (0.44) |
| CTQ-Total | 59.8 ± 20.6 | 53.3 ± 15.8 | 0.26 (0.44) |
| SSRI/SNRI use | 11 (55.0%) | 11 (47.8%) | 0.65 (0.68) |
| Beta-blocker use | 5 (25.0%) | 2 (8.7%) | 0.15 (0.36) |
| Alpha-adrenergic blocker use | 6 (30.0%) | 2 (8.7%) | 0.08 (0.24) |
| Stimulant use | 5 (25.0%) | 9 (39.1%) | 0.32 (0.44) |

<sup>a</sup> *p*-values correspond to uncorrected statistics, with false discovery rate (FDR)-corrected *p*-values shown in parentheses. For PHQ-15, SDQ-20, BDI-11, STAI, PCL-5 and CTQ, there is missing data on 3 FND participants and 1 PC participant. FND-motor, functional neurological disorder motor subtype; FND-seizure, functional neurological disorder seizure subtype; SOMS:CD, Screening for Somatoform Symptoms-7 Subscale for Conversion Disorder; SDQ-20, Somatoform Dissociation Questionnaire-20; PHQ-15 Patient Health Questionnaire-15; STAI, Spielberg State-Trait Anxiety Inventory; PCL-5, PTSD Checklist 5; CTQ, Childhood Trauma Questionnaire; SSRI/SNRI, selective serotonin reuptake inhibitor/serotonin-norepinephrine reuptake inhibitor. PHQ-15 and STAI scores were normally distributed; between-group differences were evaluated using independent sample *t*-tests. SOMS:CD, SDQ-20, BDI-II, PCL-5 and CTQ scores were not normally distributed and therefore compared using Mann-Whitney U tests. For categorical variables, chi-square tests were applied to SSRI/SNRI use and stimulant use, whereas Fisher's exact tests were used for beta-blocker use and alpha-adrenergic medication use.

Twenty-three female PCs [mean age (SD) = 33.2 ± 12.4 years] were prospectively recruited through community advertisements. PCs had a lifetime history of depression (n=19), anxiety (n=17), and/or post-traumatic stress disorder (PTSD) (n=8) (**Supplementary Table 1**). Psychiatric diagnoses were made via the Structured Clinical Interview for DSM-5 (SCID-5). Exclusion criteria for those in the PC group were the same as for those with FND, except they had no diagnosis of FND. Five additional PC female subjects were enrolled but excluded following fMRI weighted-degree outlier inspection (n=1) and quality inspection of autonomic data (n=4, see below). All subjects provided informed consent, and the Mass General Brigham Institutional Review Board approved this study.

### Procedure Overview

Participants had an in-person study visit at the Athinoula A. Martinos Center for Biomedical Imaging (Boston, MA). Upon arrival, individuals first completed a limited set of psychometric questionnaires (see *Psychometric Characterization*). Autonomic physiology at rest was then recorded using the Empatica E4 wristband under standardized conditions (see *Autonomic Data Acquisition and Preprocessing*). Following autonomic data collection, participants underwent brain MRI scanning the same day (see *MRI Acquisitions and Preprocessing*). Afterwards, participants were emailed a secure electronic link to complete the remainder of baseline study questionnaires.

### Psychometric Characterization

On the day of autonomic and rs-fMRI measurements, individuals initially completed two questionnaires: i) the Screening for Somatoform Symptoms-7 Subscale for Conversion Disorder (SOMS:CD), which is a self-report scale of FND symptoms (e.g., paralysis, impaired balance, seizures) experienced over the past week, rated on a 5-point Likert scale (Rief and Hiller, 2003); and ii) the 20-item state portion of the Spielberger State-Trait Anxiety Inventory (Spielberger, 2010). The following additional questionnaires were completed via a secure electronic link after on-site activities: 1) the Somatoform Dissociation Questionnaire-20 (SDQ-20), a 20-item scale evaluating the extent to which core FND symptoms (e.g., paralysis) were experienced over the past year on a 5-point Likert scale (Nijenhuis et al., 1996); 2) the Patient Health Questionnaire-15 (PHQ-15), a 15-item scale measuring how bothersome physical symptoms (e.g., pain, fatigue) were over the past 4 weeks on a 3-point Likert scale (Kroenke et al., 2002); 3) the 20-item trait portion of the Spielberger State-Trait Anxiety Inventory; 4) the Beck Depression Inventory (BDI-II); 5) the PTSD Checklist for DSM-5 (PCL-5); and 6) the Childhood Trauma Questionnaire (CTQ). Four participants had incomplete psychometric data (3 FND and 1 PC).

### Cardiac Autonomic Data Acquisition and Preprocessing

#### Physiological Measurement

Resting autonomic data were collected using the Empatica E4 wristband, a research-grade medical device which collects raw physiological data (CE-certified, No. 1876/MDD; www.empatica.com/research/e4/). The device measures blood volume pulse (BVP) from the photoplethysmographic (PPG) sensor, which collects data at 64 Hz through a red-green photodiode system attached to the participant’s left wrist.

Participants were instructed to turn off their mobile devices, sit upright in a comfortable position with their back supported and hands resting on their lap, and minimize unnecessary movement throughout the recording to ensure good signal quality. The Empatica E4 device was fitted on the participant’s left wrist, and data were streamed in real time to an iOS device running the E4 Connect app. After device placement, researchers visually confirmed adequate BVP signal quality and adjusted strap tension if needed to optimize sensor contact. A button press on the E4 was used to insert a timestamp marking the start of the 10-minute resting-state recording, after which research staff exited the room. The participant sat quietly and alone in a well-lit room during data collection. At the end of the 10-minute period, staff re-entered the room and inserted a second timestamp marking the end of the recording.

#### Signal Preprocessing

The raw BVP signal obtained from PPG sensors was processed using Kubios HRV Scientific software (Kubios Oy, Finland; version 4.1.2.1; Tarvainen et al., 2014). Beat-to-beat intervals were detected using the software’s built-in pulse detection algorithm (Lipponen and Tarvainen, 2019). All recordings underwent visual inspection to verify beat detection accuracy and identify segments contaminated by noise or motion artifacts; no manual correction of beat detection was performed because only artifact-free segments of data were used (see below).

To allow for physiological stabilization after placement of the E4 device, the first 60 seconds of recording were excluded for all participants. The remaining data were divided into 40–60 second segments to optimize signal quality and minimize artifact-related loss. Segments containing artifacts flagged by the Kubios software using default parameters were omitted, and a 2-beat buffer was applied before and after all detected artifacts to ensure clean IBI extraction. Following checks for artifactual segments, IBI, HR, and HRV metrics were computed for each valid segment using Kubios and subsequently exported for statistical analysis in SPSS (Version 28, IBM Corp.). Within Kubios, IBI was defined as the time (in milliseconds) between successive pulse peaks; three HRV metrics were also derived: (1) root mean square of successive differences (RMSSD) or the square root of the mean of squared differences between adjacent IBIs; (2) standard deviation of normal-to-normal intervals (SDNN) reflecting the standard deviation of all IBIs within a segment; and (3) high-frequency (HF) HRV power (0.15–0.40 Hz) via autoregressive spectral analysis of the IBI series.

For each participant, autonomic metrics were averaged across all available high-quality segments obtained within the 10-minute resting recording. Quality control procedures included visual review of raw PPG waveforms and evaluation of the physiological plausibility of the derived respiratory rate (i.e., within the high-frequency HRV band range of 0.15-0.40 Hz). Data quality characteristics are summarized in Supplementary Table 2.

### MRI Acquisition and Preprocessing

High-resolution anatomical and rs-fMRI data were acquired on a 3T scanner; full acquisition parameters are provided in the **Supplementary Materials**. Preprocessing was performed using the FMRIB Software Library (FSL v5.0.7; Oxford, UK) and MATLAB R2023a (MathWorks, Natick, MA), following pipelines previously implemented in this cohort (Westlin et al., 2025a). Anatomical (T1-weighted) preprocessing included reorientation to right-posterior-inferior (RPI) orientation, alignment to the anterior/posterior commissure plane, skull stripping, tissue segmentation into grey matter, white matter, and cerebrospinal fluid (CSF), and nonlinear registration to the 3-mm isotropic MNI152 template. Anatomical labeling and regional localization were performed using the Harvard-Oxford cortical and subcortical probabilistic atlases (Desikan et al., 2006).

Rs-fMRI preprocessing included removal of the first four volumes to allow for signal equilibration, slice-timing correction, reorientation to RPI, and within-run realignment using a six-parameter rigid-body transformation. Mean functional images were registered to individual skull-stripped T1-weighted images, followed by nonlinear transformation into MNI space. Additional preprocessing steps included spatial smoothing with a 6-mm full-width at half-maximum (FWHM) Gaussian kernel, intensity normalization, and band-pass temporal filtering (0.01–0.08 Hz). Confound regression removed 12 motion-related parameters (six affine motion estimates and their temporal derivatives), linear and quadratic trends, and five anatomical noise components each from lateral ventricular CSF and white matter masks.

Framewise displacement was computed from realignment estimates, and volumes with frame displacement > 0.5 mm were scrubbed [removed] from the data. To ensure comparable data quantity across participants, 120 volumes were retained per subject. For participants with multiple resting-state acquisitions, runs were concatenated, and the first 120 usable volumes were included in the connectivity analyses.

### Resting State Functional Connectivity Analyses

Using previously published methods (Diez et al., 2021; Westlin et al., 2025a), we investigated network centrality via whole-brain weighted-degree rsFC, which assesses each voxel’s contribution to the overall resting architecture of the brain. We first computed whole-brain rsFC matrices by extracting rs-fMRI time series from all grey matter voxels and computing Pearson correlation coefficients between each voxel pair. Negative correlations were removed due to their controversial interpretation (Qian et al., 2018). Weighted-degree was then computed as follows:

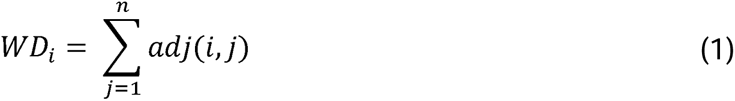

where the weighted-degree for voxel *i* (*WD_i_)* represents the sum of that voxel’s connections to all other voxels *j*. The weighted-degree of each voxel was summarized in an adjacency matrix for each participant, which was then transformed into a brain map depicting each voxel’s global rsFC.

### Statistical Analyses

#### Cardiac Autonomic Physiology and Psychometrics

Autonomic physiological metrics were compared between FND and PC cohorts using independent-samples t-tests for normally distributed metrics and Mann-Whitney U tests for non-normally distributed metrics (with false discovery rate correction applied across autonomic metrics). Associations between cardiac autonomic metrics and psychometric scores were quantified using Spearman rank correlation coefficients, again applying a false discovery rate correction. Outliers were identified using the 1.5 x interquartile range (IQR) criterion and removed on a per-metric and per-group basis prior to statistical analyses.

#### Within-Group Weighted-Degree rsFC

Primary analyses examined the relationship between weighted-degree rsFC and cardiac autonomic metrics separately within the FND and PC cohorts. For each autonomic metric, general linear models (GLMs) tested associations between weighted-degree rsFC and autonomic metrics (i.e., IBI, RMSSD, SDNN, HF-HRV) within each group. Mean HR, while reported for descriptive purposes, was not used as the physiologic metric of interest. Instead, IBI was used (inverse of HR), which has better metric qualities than HR because IBI is more linearly related to the underlying parasympathetic and sympathetic nerve activities that alter the timing of the heartbeat (Quigley and Berntson, 1996). Each model controlled for age, mean framewise displacement, SSRI/SNRI use (yes/no), and beta blocker medication use (yes/no) as covariates. Secondary (*post-hoc*) models additionally controlled for α-adrenergic blocker use (yes/no) and stimulant use (yes/no). Resulting t-statistic maps were converted to Z-statistics and corrected for multiple comparisons using cluster-wise correction via Monte Carlo simulation with 10,000 iterations, estimating the probability of observing false positive clusters at *p*<0.05.

#### Post-hoc Between-Group Analyses

To evaluate whether the correlations between weighted-degree rsFC values and indices of resting autonomic tone in participants with FND reflected regional differences as compared to values observed in PCs, we conducted *post-hoc* between-group comparisons. Specifically, for each cardiac autonomic metric that showed a significant association with weighted-degree rsFC in the FND cohort, we then compared weighted-degree values between FND and PCs within the voxels that survived all primary and *post-hoc* corrections in the main analysis. GLMs were used to compare FND vs. PCs, correcting for age and mean framewise displacement. This *post-hoc* analysis assessed whether weighted-degree rsFC values within autonomic-related regions were higher, lower, or generally the same in FND participants compared with PCs, providing additional context for interpretation of the primary results.

## RESULTS

### Demographic and Psychometric Comparisons

There were no differences between patients with FND and PCs on age, medication use (SSRI/SNRI, β-blocker, α-adrenergic blocker, or stimulant use), or scores on the BDI-II, STAI-Total, PCL-5, and CTQ-Total. However, compared with PCs, the FND cohort had increased SOMS:CD, SDQ-20, and PHQ-15 scores (**Table 1**).

### Resting Cardiac Autonomic Physiology

Mean HR, IBI, RMSSD, SDNN, and HF HRV values did not statistically differ in patients with FND compared to PCs (**Table 2**). Additionally, the cohorts were similar in terms of PPG data quality and signal characteristics from Empatica E4 recordings, including the number of segments analyzed per subject, mean segment duration, and total analyzed data duration (**Supplementary Table 2**).

**Table 2.** Comparison of Resting Heart Rate (HR) and Heart Rate Variability (HRV) Metrics Between Participants with Functional Neurological Disorder and Psychiatric Controls.

| | **Functional**<br>**Neurological Disorder**<br>**(n=20)**<br>mean ± SD | **Psychiatric**<br>**Controls**<br>**(n=23)**<br>mean ± SD | **p-value<sup>a</sup>** |
| --- | --- | --- | --- |
| HR (bpm) | 78.2 ± 10.9 | 77.9 ± 15.0 | 0.68 (0.85) |
| IBI (ms) | 782.3 ± 107.7 | 777.9 ± 129.1 | 0.91 (0.91) |
| RMSSD (ms) | 36.8 ± 23.8 | 27.6 ± 10.5 | 0.51 (0.85) |
| SDNN (ms) | 39.3 ± 20.9 | 30.5 ± 12.4 | 0.24 (0.85) |
| HF HRV (log) | 5.9 ± 1.5 | 5.6 ± 1.1 | 0.38 (0.85) |

<sup>a</sup> *p*-values correspond to uncorrected statistics, with false discovery rate (FDR)-corrected *p*-values shown in parenthesis. IBI, interbeat-interval; SDNN, standard deviation of normal-to-normal interval; RMSSD, root mean square of successive differences; HF HRV, high-frequency heart rate variability spectral power; bpm, beats per minute; ms, milliseconds; log, natural logarithm transformed values of absolute power of HF band (ms).

In both the FND and PC cohorts, cardiac autonomic metrics showed no significant correlations with psychometric variables (**Supplementary Figure 1**).

### Associations Between Cardiac Autonomic Measures and Weighted-Degree rsFC

Within the FND cohort, shorter IBIs (i.e., higher HR) were associated with increased weighted-degree rsFC in the bilateral supplementary motor area (SMA) and the right precentral / superior frontal gyri after adjusting for age, head motion, SSRI/SNRI (yes/no), and β-blocker medication use (yes/no) (**Figure 1**). These effects largely remained significant following additional *post-hoc* adjustments for α-adrenergic blocker and stimulant medication use.

**Figure 1.**
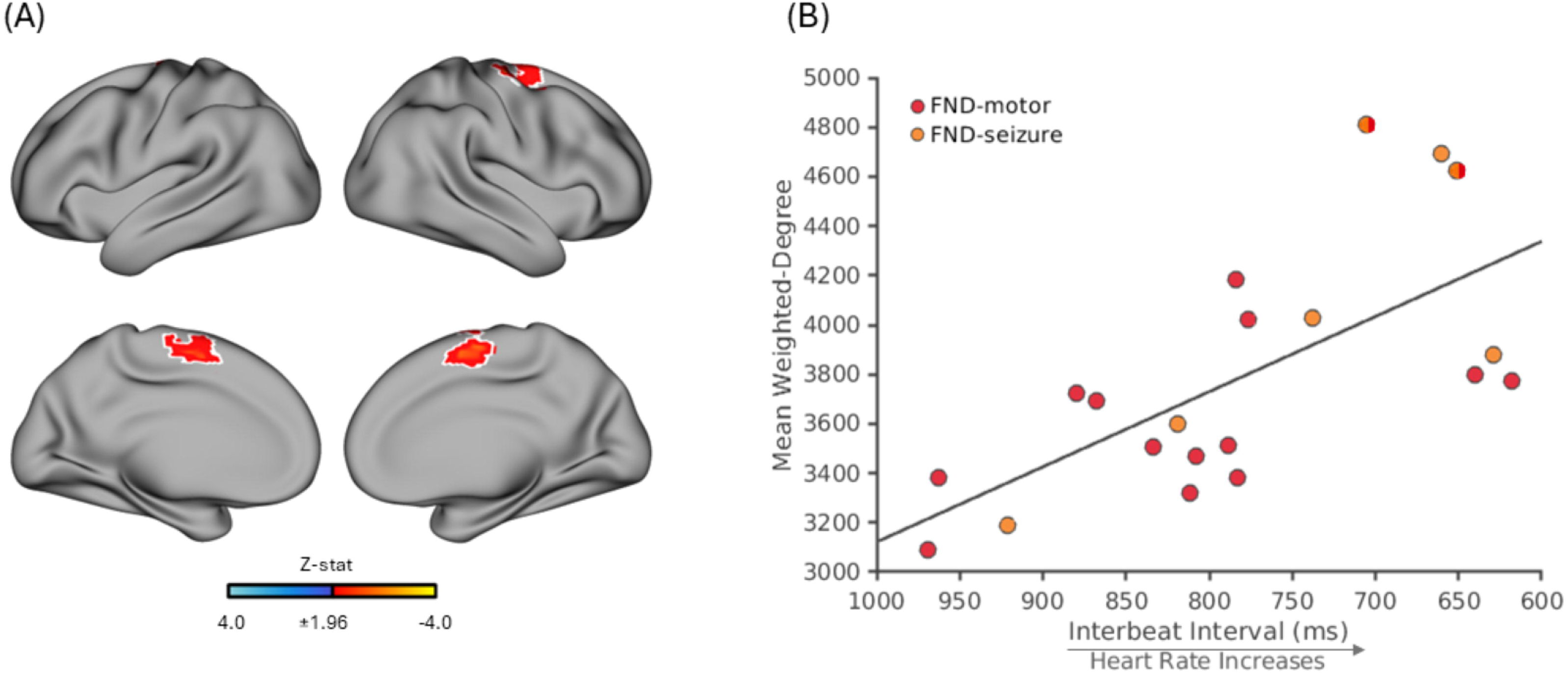
Weighted-degree resting-state functional connectivity (rsFC) correlated with interbeat interval (IBI) within female patients with functional neurological disorder (FND; n=20). **A**) Adjusting for age, head motion, SSRI/SNRI use (yes/no) and β-blocker use (yes/no), relatively shorter IBI (higher heart rate) was associated with relatively increased weighted-degree rsFC in the bilateral supplementary motor area, right precentral and superior frontal gyri. The white outline marks the subset of voxels that remain significant across *post-hoc* corrections additionally adjusting for α-adrenergic blocker use (yes/no) and stimulant medication use (yes/no). Maps show clusters defined by a voxel-wise threshold of z-statistic >1.96, corrected for multiple comparisons at the cluster level (*p* < 0.05). The volumetric brain map is shown on the cortical surface for display purposes only. **B**) Scatterplot displays mean weighted-degree values extracted from voxels within the white outlined regions versus IBI (ms). The x-axis is reversed to indicate that shorter IBI corresponds to higher heart rate (arrow). Z-stat, Z-statistic.

Within FND participants, higher time- and frequency-domain HRV metric values were consistently associated with decreased weighted-degree rsFC. Higher RMSSD (greater HRV) correlated with decreased weighted-degree in the bilateral SMA, mid-cingulate cortices, left precentral and postcentral gyri, central opercular cortex, and right orbitofrontal cortex, temporal pole, amygdala, nucleus accumbens, and putamen (**Figure 2A**). After adjusting for α-adrenergic blocker and stimulant medication use, associations remained significant within the bilateral SMA and right mid-cingulate cortex, orbitofrontal cortex, temporal pole, and amygdala regions.

**Figure 2.**
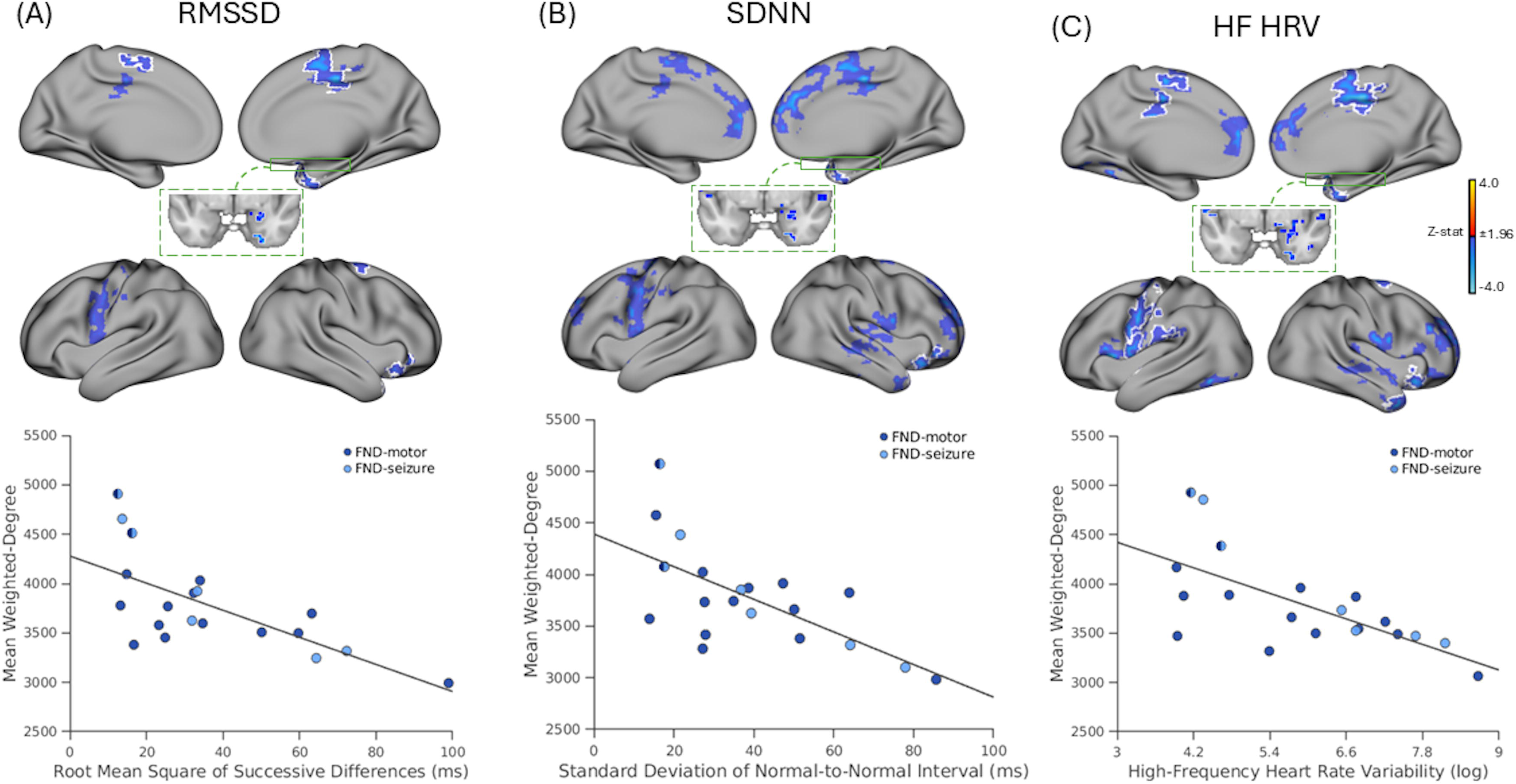
Weighted-degree resting-state functional connectivity (rsFC) correlated with time- and frequency-domain heart rate variability metrics within female patients with functional neurological disorder (FND; n=20). **A)** Adjusting for age, head motion, SSRI/SNRI use and β-blocker use, relatively higher root mean square of successive differences (RMSSD) values were associated with relative decreased weighted-degree rsFC in the bilateral supplementary motor area, mid-cingulate cortices, left precentral and postcentral gyri, central opercular cortex, and right orbitofrontal cortex, temporal pole, amygdala, nucleus accumbens, putamen (*top* panel). White outlines indicate regions that remained significant after *post-hoc* analyses additionally correcting for α-adrenergic blocker use (yes/no) and stimulant medication use (yes/no). In the *bottom* panel, the scatterplot displays mean weighted-degree values extracted from voxels within the white outlined regions versus RMSSD (ms). **B)** Adjusting for age, head motion, SSRI/SNRI use and β-blocker use, relatively higher standard deviation of normal-to-normal intervals (SDNN) was associated with relative decreased weighted-degree rsFC in the bilateral supplementary motor area, anterior and mid-cingulate cortices, paracingulate gyri, medial prefrontal cortices, frontal poles, central opercular cortices, left precentral and superior frontal gyri, and right orbitofrontal cortex, insula and insular-opercular cortex, middle temporal gyrus, temporal pole, and amygdala (*top* panel). White outlines indicate regions that remained significant after *post-hoc* corrections additionally adjusting for α-adrenergic blocker use (yes/no) and stimulant medication use (yes/no). In the *bottom* panel, the scatterplot displays mean weighted-degree values extracted from voxels within the white outlined regions this versus SDNN (ms). **C)** Adjusting for age, head motion, SSRI/SNRI use and β-blocker use, relatively higher values of high frequency heart rate variability (HF HRV) were associated with relative decreased weighted-degree rsFC in the bilateral supplementary motor area, anterior and mid-cingulate cortices, paracingulate gyri, medial prefrontal cortices, central opercular cortices, left precentral, postcentral, and lingual gyri, and right orbitofrontal cortex, inferior frontal gyrus, frontal pole, insula, middle temporal gyrus, temporal pole, amygdala, thalamus, and putamen (*top* panel). White outlines indicate regions that remained significant after additional post-hoc corrections for α-adrenergic blocker use and stimulant medication use. In the *bottom* panel, the scatterplot displays mean weighted-degree values extracted from voxels within the white outlined regions versus HF HRV (ln-ms²). Maps show clusters defined by a voxel-wise threshold of z-statistic >1.96; corrected for multiple comparisons at the cluster level (*p*<0.05). Volumetric brain maps are visualized on the cortical surface for display purposes only. Z-stat, Z-statistic; RMSSD, root mean square of successive differences; SDNN, standard deviation of normal-to-normal intervals; HF HRV, high frequency heart rate variability.

Similarly, higher SDNN (also reflective of greater HRV) was associated with decreased weighted-degree rsFC in the bilateral SMA, anterior and mid-cingulate cortices, paracingulate gyri, medial prefrontal cortices, frontal poles, central opercular cortices, left precentral, and superior frontal gyri, and right orbitofrontal cortex, insula and insular-opercular cortex, superior/middle temporal gyrus, temporal pole and the amygdala (**Figure 2B**). After adjusting for α-adrenergic blocker and stimulant medication use, associations remained significant within the right orbitofrontal cortex, temporal pole and amygdala.

Finally, higher HF HRV was associated with decreased weighted-degree rsFC across the bilateral SMA, anterior and mid-cingulate cortices, paracingulate gyri, medial prefrontal cortices, central opercular cortices, left precentral, postcentral, and lingual gyri, and right orbitofrontal cortex, inferior frontal gyrus, frontal pole, insula, middle temporal gyrus, and temporal pole regions, as well as the right amygdala, thalamus, and putamen (**Figure 2C**). After adjusting for α-adrenergic blocker and stimulant medication use, associations remained significant within the bilateral SMA, the right amygdala, temporal pole, insula, orbitofrontal cortex, and the left precentral, postcentral, and lingual gyri.

Notably, within FND participants all three HRV metrics showed significant associations with weighted-degree rsFC in the right amygdala, temporal pole, and orbitofrontal cortex that held across all *post-hoc* adjustments. In addition, both RMSSD and HF HRV showed consistent associations with weighted-degree in the bilateral SMA across all *post-hoc* adjustments. See **Supplementary Figure 2** for all significant rsFC-autonomic relationships in females with FND.

Within PCs, cardiac autonomic indices–rsFC associations were more spatially restricted. Higher RMSSD was associated with decreased weighted-degree rsFC in only the bilateral anterior/mid-cingulate cortices, and these effects remained significant after adjusting for α-adrenergic blocker and stimulant medication use (**Figure 3A**). Similarly, higher HF HRV was associated with decreased weighted-degree rsFC within the bilateral anterior/mid-cingulate cortices and SMA, with associations remaining significant after both medication adjustments (**Figure 3B**). No significant relationships were observed for IBI or SDNN. See **Supplementary Figure 3** for all significant rsFC-autonomic relationships in female PCs.

**Figure 3.**
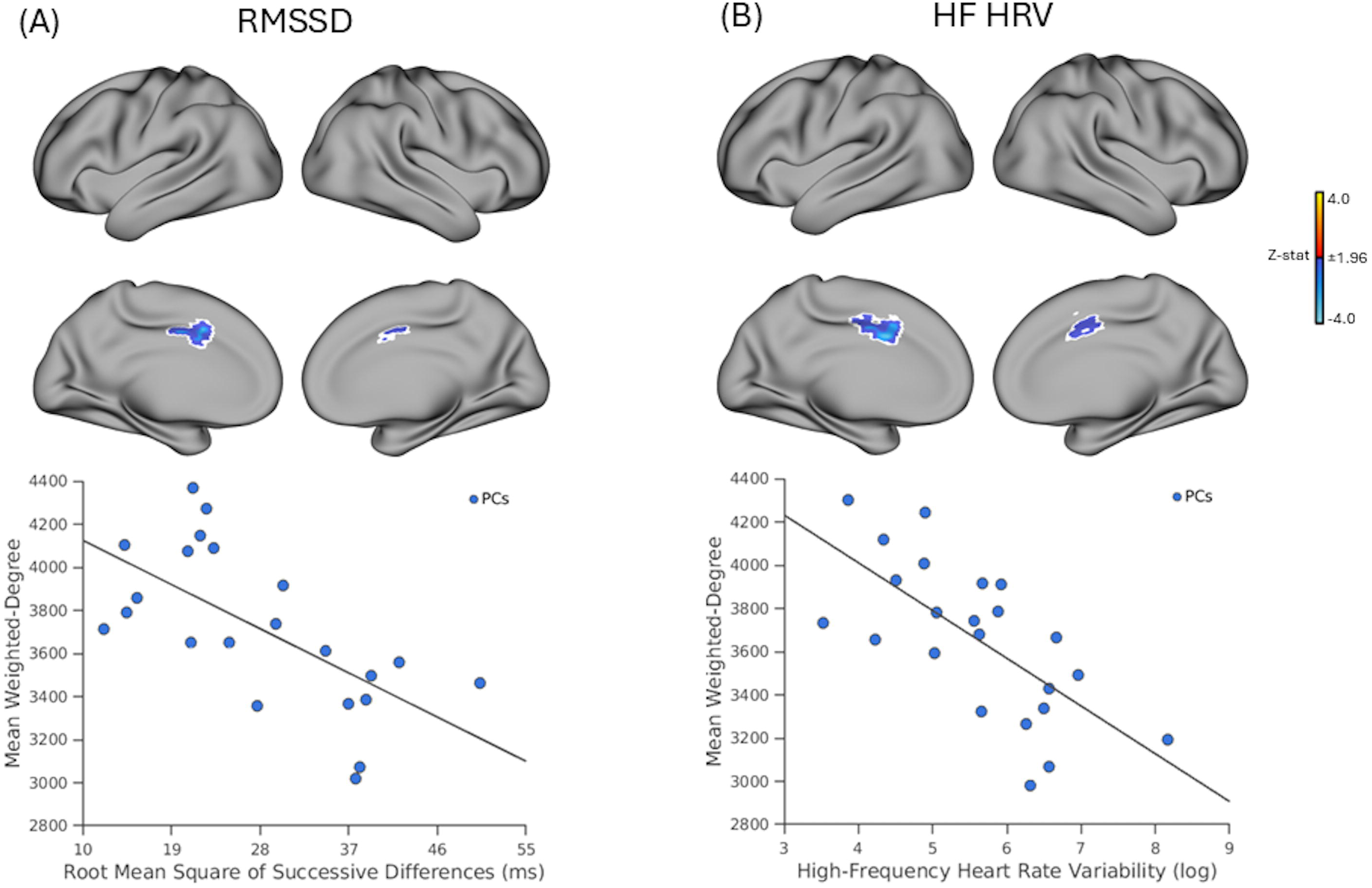
Weighted-degree resting-state functional connectivity (rsFC) correlated with time- and frequency-domain HRV metrics within female psychiatric controls (n=22 for RMSSD and n=23 for HF HRV). A) Adjusting for age, head motion, SSRI/SNRI use and β-blocker use, relatively higher root mean square of successive difference (RMSSD) values were associated with relatively decreased weighted-degree rsFC in the bilateral anterior/mid-cingulate cortices (*top* panel). In the *bottom* panel, the scatterplot displays mean weighted-degree values extracted from voxels within the white outlined regions versus RMSSD (ms). B) Adjusting for age, head motion, SSRI/SNRI use and β-blocker use, relatively higher high-frequency heart rate variability (HF HRV) was associated with relative decreased weighted-degree rsFC in the bilateral anterior/mid-cingulate cortices and the supplementary motor area (*top* panel). In the *bottom* panel, the scatterplot displays mean weighted-degree values extracted from voxels within the white outlined regions versus HF HRV (ln-ms²). For *top* panels (A and B), the white outline marks the subset of voxels that remain significant across *post-hoc* corrections additionally adjusting for α-adrenergic blocker use (yes/no) and stimulant medication use (yes/no). Maps show clusters defined by a voxel-wise threshold of *z* > 1.96, corrected for multiple comparisons at the cluster level (*p* < 0.05). Volumetric brain maps are visualized on the cortical surface for display purposes only. Z-stat, Z-statistic; RMSSD, root mean square of successive differences; SDNN, standard deviation of normal-to-normal intervals.

### Between-Group Weighted-Degree Comparison within Cardiac Autonomic-Related Regions

When investigating potential group-level [FND vs. PC] differences in the brain areas that showed positive correlations between cardiac autonomic metrics and weighted-degree rsFC in patients with FND, *post-hoc* comparisons with the PC cohort revealed subtle differences, characterized primarily by modestly increased and more variable rsFC values in the FND cohort. Specifically, patients with FND showed increased weighted-degree rsFC relative to PCs within the right SMA across IBI-, RMSSD-, and HF HRV-related masks, and within the right amygdala across RMSSD-, SDNN-, and HF HRV-related masks (**Figure 4).**

**Figure 4.**
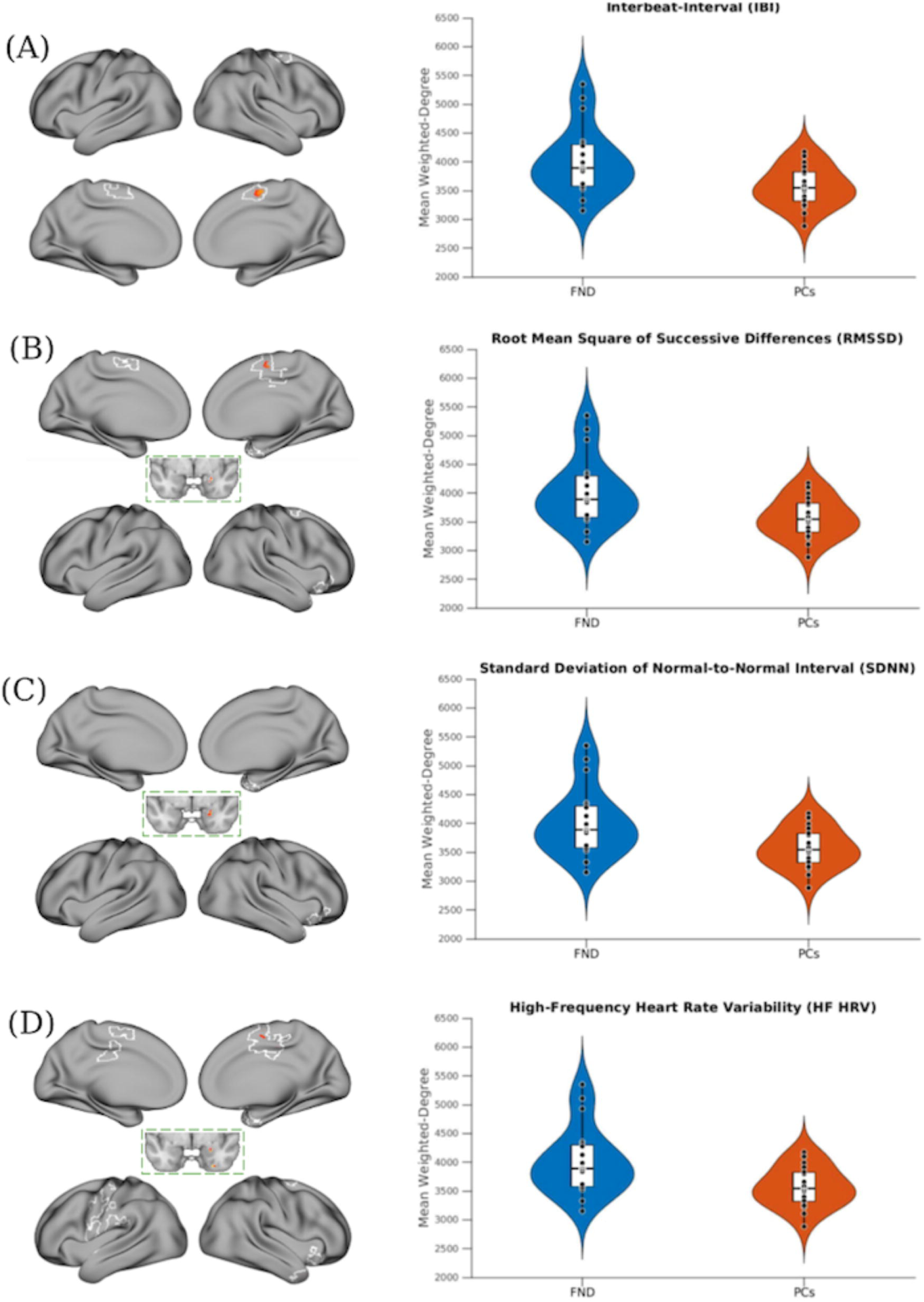
Between-group *post-hoc* comparison of mean weighted-degree resting-state functional connectivity (rsFC) within IBI, and time- and frequency-domain heart rate variability (HRV)-metrics correlated masks. A-D) Maps on the left show voxels where weighted-degree rsFC differed between patients with functional neurological disorder (FND) and psychiatric controls (PCs), restricted to the subset of voxels that remained significant across all *post-hoc* corrections in the IBI and HRV analyses (Figure 1 and Figure 2). Violin plots on the right column show the distribution of mean weighted-degree values extracted from each IBI/HRV metric-correlated masks that remained significant after all *post-hoc* adjustments (white outline in Figures 1 and 2). FND participants exhibited increased weighted-degree rsFC values within these regions compared to PCs. For all display items, individual dots represent participants; white boxes indicate interquartile ranges and medians. Maps show clusters defined by a voxel-wise threshold of z-statistic >1.96, corrected for multiple comparisons at the cluster level (*p* < 0.05). The volumetric brain map is shown on the cortical surface for display purposes only. RMSSD, root mean square of successive differences; SDNN, standard deviation of normal-to-normal intervals; HF HRV, high frequency heart rate variability.

## DISCUSSION

In this pilot study, resting cardiac autonomic metrics did not distinguish female patients with FND from age-matched female PCs. Cardiac autonomic metrics also were not significantly associated with self-reported FND symptom severity, somatic symptom burden, affective symptoms, or the magnitude of adverse childhood experiences. Nonetheless, shorter IBIs (reflecting higher HR) in patients with FND correlated with relatively greater weighted-degree rsFC in the bilateral SMA and primary motor regions. Greater HRV (higher RMSSD, SDNN, HF HRV) was also linked to reduced weighted-degree rsFC, primarily within bilateral SMA, cingulate, temporal pole, and right orbitofrontal cortex, insula, and amygdala. *Post-hoc* analyses further clarified that individuals with FND exhibited slightly increased and more variable weighted-degree values in the right SMA and right amygdala compared to PCs – indicating that these areas had a greater [albeit modestly different] contribution to overall functional brain architecture at rest in the FND cohort than in PCs. It should also be noted that in most areas showing relationships between rsFC and cardiac autonomic metrics at rest in patients with FND, the weighted-degree values in those brain regions were comparable to PCs. Collectively, these findings suggest that individual-subject cardiac autonomic profiles in FND correlate with the brain organization of multiple rsFC networks previously implicated in the pathophysiology of FND (Drane et al., 2020; Yeo et al., 2011).

In the FND-related autonomic literature, several studies reported elevated resting HR and lower HRV in patients with FND compared to HCs (Kozlowska et al., 2015; Maurer et al., 2016), whereas others have found no group-level differences in these same metrics (Demartini et al., 2016; Ponnusamy et al., 2012). However, many of these prior studies did not fully account for comorbid depression, anxiety, trauma history, and psychotropic medication use, each of which can impact cardiac autonomic metrics (Alvares et al., 2016; Chalmers et al., 2014; Kemp et al., 2010). In this current study, our controls were well-matched PCs, and we preliminarily show that there may be no categorically distinguishing cardiac autonomic profile at rest in adult females with FND at a group-level when compared to a psychiatric (rather than healthy) control group. The absence of between-group differences here does not imply an absence of autonomic relevance; rather, our findings suggest that autonomic features are common in individuals with psychiatric and neuropsychiatric conditions, and may also highlight substantial biological heterogeneity within FND – emphasizing the need to take individual differences into account (Diez et al., 2019; Perez et al., 2017). Notably, similar heterogeneous observations have been found when investigating potential performance differences on a heartbeat tracking task (assessing the tendency to report the timing of one’s heartbeats with accuracy), with some FND cohort studies showing between-group differences in relation to HCs (Koreki et al., 2020; Ricciardi et al., 2014) and others showing similar performance across groups (Jungilligens et al., 2020; Pick et al., 2020; Sojka et al., 2021). Relatedly, differences in heartbeat tracking accuracy across patients with FND related to individual differences in cingulo-insular activation (Sojka et al., 2025). These results underscore that various physiological measurements in FND may not be best conceptualized as standalone biomarkers, but rather by *how* physiological measurements (e.g., cardiac autonomic metrics at rest) relate to other biological systems—such as functional brain network architecture.

Across the IBI and HRV within-group analyses in patients with FND, the SMA emerged as a consistently implicated region, with shorter IBI [higher HR] associated with elevated rsFC weighted-degree values and higher HRV associated with reduced rsFC values. These observations are noteworthy given that several fMRI studies have identified a role for the SMA and its connections in the pathophysiology of FND. For example, task-based fMRI studies have shown increased SMA activation during recall of affectively charged autobiographical events (Aybek et al., 2014), increased SMA recruitment during exposure to normatively negative emotional stimuli (Aybek et al., 2015), and abnormal SMA activity during motor preparation even when no movement was performed (Voon et al., 2011). In a pilot real-time fMRI study, increases in subjective agency in patients with FND also were associated with upregulation of SMA and right temporoparietal junction activity (Müller et al., 2025). Prior work from our group has also shown that rsFC relationships between the anterior insula and the SMA relate to individual differences in functional neurological symptom severity (Diez et al., 2019). SMA rsFC profiles, as measured using Blood-Oxygen-Level-Dependent (BOLD) signal variability, have also shown prognostic relevance in patients with FND (Schneider et al., 2024). Neurobiologically, the SMA is a premotor hub theorized to be involved in internally-generated action planning and motor initiation, and it is functionally connected with the somatomotor and salience networks (Hallett, 2022). As such, Voon et al (2011) proposed that during states of heightened physiological arousal there may be a disruption in the action-selection process that is in part mediated through the SMA (Voon et al., 2011). In relation to the findings of this study, it is important to highlight that the SMA is a cortical component of the CAN (Beissner et al., 2013; Valenza et al., 2019). Given the results of this present study, we speculate that the SMA may be a hub through which cardiac autonomic metrics influence motor expression patterns in patients with FND.

Beyond the SMA, individual differences in weighted-degree relationships to HRV metrics were also observed in the right amygdala, anterior insula and lateral orbitofrontal cortex. Specifically, higher HRV correlated with reduced centrality of these brain areas in the overall functional architecture; conversely, those patients with FND *and* relatively lower HRV indices showed increased amygdala, anterior insula and orbitofrontal cortex influence on their overall functional brain architecture. These brain regions overlap in their contributions to the CAN, the allostatic-interoceptive network, and several canonical resting-state functional networks (i.e., salience and limbic networks) (Kleckner et al., 2017; Quadt et al., 2022; Yeo et al., 2011). Given that motor control areas [including but not limited to the SMA] have shown increased functional connectivity to the amygdala and insula across task and rs-fMRI studies in patients with FND (Aybek et al., 2015, 2014; Diez et al., 2019; Li et al., 2015b; Voon et al., 2011), the findings of this present work suggest that individual differences in cardiac autonomic metrics could relate to the role of the amygdala and anterior insula in overall functional brain organization. Additionally, while we did not identify correlations between cardiac autonomic metrics at rest and childhood maltreatment burden [potentially due to modest sample size], such early-life factors may influence the autonomic nervous system over the long-term – including promoting tendencies towards having lower HRV (Stevens et al., 2023; Stürmer et al., 2025). This literature is noteworthy given “dose-dependent relationships” previously reported between the magnitude of childhood physical abuse burden and the rsFC strength relationships between the amygdala and insula to the primary motor cortex (Diez et al., 2021). Taken together, these findings are consistent with the notion that in some patients with FND, hubs within the CAN, the allostatic-interoceptive network, and the salience network show altered contributions to the overall functional network architecture.

The inclusion of a psychiatric comparison group also provides important insight into the biological heterogeneity of FND. While associations between weighted-degree and cardiac autonomic metrics were present in both groups, they were more spatially extensive in FND, whereas PCs showed a more limited pattern centered mainly on the anterior/mid-cingulate cortices. *Post-hoc* analyses further support this interpretation: within autonomic-associated regions, patients with FND exhibited slightly higher and more variable weighted-degree values in the right SMA and right amygdala compared to PCs. On inspection of the data, brain-autonomic relationships in patients with FND appear more robust and widespread in part because of the larger between-subject heterogeneity observed in this population.

Several limitations are important when interpreting these findings. First, the sample size is modest, and as such, the results should be considered preliminary. Additional research is also needed to explore sex differences in brain-autonomic relationships in patients with FND, as well as the need to investigate potential differences across FND phenotypes. Furthermore, cardiac autonomic metrics were assessed at rest; while this approach reduces task-related confounds, it does not capture dynamic autonomic responsivity in the context of provocation. Finally, the cross-sectional study design cannot determine whether cardiac autonomic–brain network associations represent a predisposing vulnerability, a perpetuating factor, and/or compensatory mechanisms (among many possibilities). Large-scale multimodal brain imaging efforts, along with a more comprehensive sampling of autonomic parameters (e.g., characterization of autonomic variables beyond cardiac metrics), will be important to replicate and extend the findings of this initial study (Roberts et al., 2020, 2012).

In summary, this pilot study provides early-phase evidence that individual differences in cardiac autonomic metrics relate to large-scale brain network architecture in FND. Moving forward, multimodal physiological sampling — including the dual acquisition of brain imaging and peripheral autonomic measures — may provide a more complete and nuanced understanding of the neurobiology of FND.

## Acknowledgments

We thank all the research subjects for their study participation.

## Data Availability

For qualified researchers, analysis code and de-identified data pertaining to study results can be made available upon reasonable request and following approval by the local internal review board.

## References

1. Allendorfer, J.B., Nenert, R., Hernando, K.A., DeWolfe, J.L., Pati, S., Thomas, A.E., Billeaud, N., Martin, R.C., Szaflarski, J.P., 2019. FMRI response to acute psychological stress differentiates patients with psychogenic non-epileptic seizures from healthy controls - A biochemical and neuroimaging biomarker study. NeuroImage Clin. 24, 101967. 10.1016/j.nicl.2019.101967

2. Alvares, G.A., Quintana, D.S., Hickie, I.B., Guastella, A.J., 2016. Autonomic nervous system dysfunction in psychiatric disorders and the impact of psychotropic medications: a systematic review and meta-analysis. J. Psychiatry Neurosci. JPN 41, 89–104. 10.1503/jpn.140217

3. Anderson, K.E., Gruber-Baldini, A.L., Vaughan, C.G., Reich, S.G., Fishman, P.S., Weiner, W.J., Shulman, L.M., 2007. Impact of psychogenic movement disorders versus Parkinson’s on disability, quality of life, and psychopathology. Mov. Disord. Off. J. Mov. Disord. Soc. 22, 2204–2209. 10.1002/mds.21687

4. Aybek, S., Nicholson, T.R., O’Daly, O., Zelaya, F., Kanaan, R.A., David, A.S., 2015. Emotion-motion interactions in conversion disorder: an FMRI study. PloS One 10, e0123273. 10.1371/journal.pone.0123273

5. Aybek, S., Nicholson, T.R., Zelaya, F., O’Daly, O.G., Craig, T.J., David, A.S., Kanaan, R.A., 2014. Neural correlates of recall of life events in conversion disorder. JAMA Psychiatry 71, 52–60. 10.1001/jamapsychiatry.2013.2842

6. Aybek, S., Perez, D.L., 2022. Diagnosis and management of functional neurological disorder. BMJ 376, o64. 10.1136/bmj.o64

7. Baizabal-Carvallo, J.F., Hallett, M., Jankovic, J., 2019. Pathogenesis and pathophysiology of functional (psychogenic) movement disorders. Neurobiol. Dis. 127, 32–44. 10.1016/j.nbd.2019.02.013

8. Bakvis, P., Roelofs, K., Kuyk, J., Edelbroek, P.M., Swinkels, W.A.M., Spinhoven, P., 2009. Trauma, stress, and preconscious threat processing in patients with psychogenic nonepileptic seizures. Epilepsia 50, 1001–1011. 10.1111/j.1528-1167.2008.01862.x

9. Beissner, F., Meissner, K., Bär, K.-J., Napadow, V., 2013. The autonomic brain: an activation likelihood estimation meta-analysis for central processing of autonomic function. J. Neurosci. Off. J. Soc. Neurosci. 33, 10503–10511. 10.1523/JNEUROSCI.1103-13.2013

10. Benarroch, E.E., 1993. The central autonomic network: functional organization, dysfunction, and perspective. Mayo Clin. Proc. 68, 988–1001. 10.1016/s0025-6196(12)62272-1

11. Butler, M., Shipston-Sharman, O., Seynaeve, M., Bao, J., Pick, S., Bradley-Westguard, A., Ilola, E., Mildon, B., Golder, D., Rucker, J., Stone, J., Nicholson, T., 2021. International online survey of 1048 individuals with functional neurological disorder. Eur. J. Neurol. 28, 3591–3602. 10.1111/ene.15018

12. Cechetto, D.F., Shoemaker, J.K., 2009. Functional neuroanatomy of autonomic regulation. NeuroImage 47, 795–803. 10.1016/j.neuroimage.2009.05.024

13. Chalmers, J.A., Quintana, D.S., Abbott, M.J.-A., Kemp, A.H., 2014. Anxiety Disorders are Associated with Reduced Heart Rate Variability: A Meta-Analysis. Front. Psychiatry 5, 80. 10.3389/fpsyt.2014.00080

14. Demartini, B., Goeta, D., Barbieri, V., Ricciardi, L., Canevini, M.P., Turner, K., D’Agostino, A., Romito, L., Gambini, O., 2016a. Psychogenic non-epileptic seizures and functional motor symptoms: A common phenomenology? J. Neurol. Sci. 368, 49–54. 10.1016/j.jns.2016.06.045

15. Demartini, B., Goeta, D., Barbieri, V., Ricciardi, L., Canevini, M.P., Turner, K., D’Agostino, A., Romito, L., Gambini, O., 2016b. Psychogenic non-epileptic seizures and functional motor symptoms: A common phenomenology? J. Neurol. Sci. 368, 49–54. 10.1016/j.jns.2016.06.045

16. Demartini, B., Nisticò, V., Edwards, M.J., Gambini, O., Priori, A., 2021. The pathophysiology of functional movement disorders. Neurosci. Biobehav. Rev. 120, 387–400. 10.1016/j.neubiorev.2020.10.019

17. Desikan, R.S., Ségonne, F., Fischl, B., Quinn, B.T., Dickerson, B.C., Blacker, D., Buckner, R.L., Dale, A.M., Maguire, R.P., Hyman, B.T., Albert, M.S., Killiany, R.J., 2006. An automated labeling system for subdividing the human cerebral cortex on MRI scans into gyral based regions of interest. NeuroImage 31, 968–980. 10.1016/j.neuroimage.2006.01.021

18. Diez, I., Larson, A.G., Nakhate, V., Dunn, E.C., Fricchione, G.L., Nicholson, T.R., Sepulcre, J., Perez, D.L., 2021. Early-life trauma endophenotypes and brain circuit-gene expression relationships in functional neurological (conversion) disorder. Mol. Psychiatry 26, 3817–3828. 10.1038/s41380-020-0665-0

19. Diez, I., Ortiz-Terán, L., Williams, B., Jalilianhasanpour, R., Ospina, J.P., Dickerson, B.C., Keshavan, M.S., LaFrance, W.C., Sepulcre, J., Perez, D.L., 2019. Corticolimbic fast-tracking: enhanced multimodal integration in functional neurological disorder. J. Neurol. Neurosurg. Psychiatry 90, 929–938. 10.1136/jnnp-2018-319657

20. Drane, D.L., Fani, N., Hallett, M., Khalsa, S.S., Perez, D.L., Roberts, N.A., 2020. A framework for understanding the pathophysiology of functional neurological disorder. CNS Spectr. 1–7. 10.1017/S1092852920001789

21. Finkelstein, S.A., Diamond, C., Carson, A., Stone, J., 2025. Incidence and prevalence of functional neurological disorder: a systematic review. J. Neurol. Neurosurg. Psychiatry 96, 383–395. 10.1136/jnnp-2024-334767

22. Goetz, A.R., Lee, H.J., Cougle, J.R., 2013. The association between health anxiety and disgust reactions in a contamination-based behavioral approach task. Anxiety Stress Coping 26, 431–46. 10.1080/10615806.2012.684241

23. Goldstein, L.H., Mellers, J.D.C., 2006. Ictal symptoms of anxiety, avoidance behaviour, and dissociation in patients with dissociative seizures. J. Neurol. Neurosurg. Psychiatry 77, 616–621. 10.1136/jnnp.2005.066878

24. Gray, C., Calderbank, A., Adewusi, J., Hughes, R., Reuber, M., 2020. Symptoms of posttraumatic stress disorder in patients with functional neurological symptom disorder. J. Psychosom. Res. 129, 109907. 10.1016/j.jpsychores.2019.109907

25. Hallett, M., 2022. Free Will, Emotions and Agency: Pathophysiology of Functional Movement Disorder, in: LaFaver, K., Maurer, C.W., Nicholson, T.R., Perez, D.L. (Eds.), Functional Movement Disorder, Current Clinical Neurology. Springer International Publishing, Cham, pp. 13–26. 10.1007/978-3-030-86495-8_2

26. Hallett, M., Aybek, S., Dworetzky, B.A., McWhirter, L., Staab, J.P., Stone, J., 2022. Functional neurological disorder: new subtypes and shared mechanisms. Lancet Neurol. 21, 537–550. 10.1016/S1474-4422(21)00422-1

27. Homayoun, M., Lo, A.P.-K., Sinclair, B., Nazem-Zadeh, M.-R., Neal, A., Winton-Brown, T., Kwan, P., Kanaan, R.A., 2025. Neuroimaging abnormalities in functional/dissociative seizures: a meta-analysis. Neurosci. Biobehav. Rev. 180, 106439. 10.1016/j.neubiorev.2025.106439

28. Indranada, A.M., Mullen, S.A., Wong, M.J., D’Souza, W.J., Kanaan, R.A.A., 2019. Preictal autonomic dynamics in psychogenic nonepileptic seizures. Epilepsy Behav. EB 92, 206–212. 10.1016/j.yebeh.2018.12.026

29. Jones, B., Reuber, M., Norman, P., 2016. Correlates of health-related quality of life in adults with psychogenic nonepileptic seizures: A systematic review. Epilepsia 57, 171–181. 10.1111/epi.13268

30. Jungilligens, J., Paredes-Echeverri, S., Popkirov, S., Barrett, L.F., Perez, D.L., 2022. A new science of emotion: implications for functional neurological disorder. Brain J. Neurol. 145, 2648–2663. 10.1093/brain/awac204

31. Jungilligens, J., Wellmer, J., Schlegel, U., Kessler, H., Axmacher, N., Popkirov, S., 2020. Impaired emotional and behavioural awareness and control in patients with dissociative seizures. Psychol. Med. 50, 2731–2739. 10.1017/S0033291719002861

32. Kemp, A.H., Quintana, D.S., Gray, M.A., Felmingham, K.L., Brown, K., Gatt, J.M., 2010. Impact of depression and antidepressant treatment on heart rate variability: a review and meta-analysis. Biol. Psychiatry 67, 1067–1074. 10.1016/j.biopsych.2009.12.012

33. Kleckner, I.R., Zhang, J., Touroutoglou, A., Chanes, L., Xia, C., Simmons, W.K., Quigley, K.S., Dickerson, B.C., Barrett, L.F., 2017. Evidence for a Large-Scale Brain System Supporting Allostasis and Interoception in Humans. Nat. Hum. Behav. 1, 0069. 10.1038/s41562-017-0069

34. Koreki, A., Garfkinel, S.N., Mula, M., Agrawal, N., Cope, S., Eilon, T., Gould Van Praag, C., Critchley, H.D., Edwards, M., Yogarajah, M., 2020. Trait and state interoceptive abnormalities are associated with dissociation and seizure frequency in patients with functional seizures. Epilepsia 61, 1156–1165. 10.1111/epi.16532

35. Kozlowska, K., Palmer, D.M., Brown, K.J., McLean, L., Scher, S., Gevirtz, R., Chudleigh, C., Williams, L.M., 2015a. Reduction of autonomic regulation in children and adolescents with conversion disorders. Psychosom. Med. 77, 356–370. 10.1097/PSY.0000000000000184

36. Kozlowska, K., Palmer, D.M., Brown, K.J., McLean, L., Scher, S., Gevirtz, R., Chudleigh, C., Williams, L.M., 2015b. Reduction of autonomic regulation in children and adolescents with conversion disorders. Psychosom. Med. 77, 356–370. 10.1097/PSY.0000000000000184

37. Kozlowska, K., Rampersad, R., Cruz, C., Shah, U., Chudleigh, C., Soe, S., Gill, D., Scher, S., Carrive, P., 2017. The respiratory control of carbon dioxide in children and adolescents referred for treatment of psychogenic non-epileptic seizures. Eur. Child Adolesc. Psychiatry 26, 1207–1217. 10.1007/s00787-017-0976-0

38. Kozlowska, K., Spooner, C.J., Palmer, D.M., Harris, A., Korgaonkar, M.S., Scher, S., Williams, L.M., 2018. “Motoring in idle”: The default mode and somatomotor networks are overactive in children and adolescents with functional neurological symptoms. NeuroImage Clin. 18, 730–743. 10.1016/j.nicl.2018.02.003

39. Kroenke, K., Spitzer, R.L., Williams, J.B.W., 2002. The PHQ-15: validity of a new measure for evaluating the severity of somatic symptoms. Psychosom. Med. 64, 258–266. 10.1097/00006842-200203000-00008

40. LaFrance, W.C., Baker, G.A., Duncan, R., Goldstein, L.H., Reuber, M., 2013a. Minimum requirements for the diagnosis of psychogenic nonepileptic seizures: a staged approach: a report from the International League Against Epilepsy Nonepileptic Seizures Task Force. Epilepsia 54, 2005–2018. 10.1111/epi.12356

41. LaFrance, W.C., Baker, G.A., Duncan, R., Goldstein, L.H., Reuber, M., 2013b. Minimum requirements for the diagnosis of psychogenic nonepileptic seizures: a staged approach: a report from the International League Against Epilepsy Nonepileptic Seizures Task Force. Epilepsia 54, 2005–2018. 10.1111/epi.12356

42. Li, R., Li, Y., An, D., Gong, Q., Zhou, D., Chen, H., 2015a. Altered regional activity and inter-regional functional connectivity in psychogenic non-epileptic seizures. Sci. Rep. 5, 11635. 10.1038/srep11635

43. Li, R., Liu, K., Ma, X., Li, Z., Duan, X., An, D., Gong, Q., Zhou, D., Chen, H., 2015b. Altered Functional Connectivity Patterns of the Insular Subregions in Psychogenic Nonepileptic Seizures. Brain Topogr. 28, 636–645. 10.1007/s10548-014-0413-3

44. Lipponen, J.A., Tarvainen, M.P., 2019. A robust algorithm for heart rate variability time series artefact correction using novel beat classification. J. Med. Eng. Technol. 43, 173–181. 10.1080/03091902.2019.1640306

45. Maurer, C.W., Liu, V.D., LaFaver, K., Ameli, R., Wu, T., Toledo, R., Epstein, S.A., Hallett, M., 2016a. Impaired resting vagal tone in patients with functional movement disorders. Parkinsonism Relat. Disord. 30, 18–22. 10.1016/j.parkreldis.2016.06.009

46. Maurer, C.W., Liu, V.D., LaFaver, K., Ameli, R., Wu, T., Toledo, R., Epstein, S.A., Hallett, M., 2016b. Impaired resting vagal tone in patients with functional movement disorders. Parkinsonism Relat. Disord. 30, 18–22. 10.1016/j.parkreldis.2016.06.009

47. Mavroudis, I., Kazis, D., Kamal, F.Z., Gurzu, I.-L., Ciobica, A., Pădurariu, M., Novac, B., Iordache, A., 2024. Understanding Functional Neurological Disorder: Recent Insights and Diagnostic Challenges. Int. J. Mol. Sci. 25, 4470. 10.3390/ijms25084470

48. McKenzie, P.S., Oto, M., Graham, C.D., Duncan, R., 2011. Do patients whose psychogenic non-epileptic seizures resolve, “replace” them with other medically unexplained symptoms? Medically unexplained symptoms arising after a diagnosis of psychogenic non-epileptic seizures. J. Neurol. Neurosurg. Psychiatry 82, 967–969. 10.1136/jnnp.2010.231886

49. Morris, L.S., To, B., Baek, K., Chang-Webb, Y.-C., Mitchell, S., Strelchuk, D., Mikheenko, Y., Phillips, W., Zandi, M., Jenaway, A., Walsh, C., Voon, V., 2017. Disrupted avoidance learning in functional neurological disorder: Implications for harm avoidance theories. NeuroImage Clin. 16, 286–294. 10.1016/j.nicl.2017.08.007

50. Müller, E., Loukas, S., Häuselmann, S., Concetti, C., Van De Ville, D., Gninenko, N., Aybek, S., 2025. Modulating the sense of agency in functional neurological disorder using real-time fMRI neurofeedback: a proof-of-concept study. NeuroImage Clin. 48, 103899. 10.1016/j.nicl.2025.103899

51. Nijenhuis, E.R., Spinhoven, P., Van Dyck, R., Van der Hart, O., Vanderlinden, J., 1996. The development and psychometric characteristics of the Somatoform Dissociation Questionnaire (SDQ-20). J. Nerv. Ment. Dis. 184, 688–694. 10.1097/00005053-199611000-00006

52. O’Mahony, B., Nielsen, G., Baxendale, S., Edwards, M.J., Yogarajah, M., 2023. Economic Cost of Functional Neurologic Disorders: A Systematic Review. Neurology 101, e202–e214. 10.1212/WNL.0000000000207388

53. Paredes-Echeverri, S., Maggio, J., Bègue, I., Pick, S., Nicholson, T.R., Perez, D.L., 2022 Autonomic, Endocrine, and Inflammation Profiles in Functional Neurological Disorder: A Systematic Review and Meta-Analysis. J Neuropsychiatry Clin Neurosci. 34(1):30–43. 10.1176/appi.neuropsych.21010025

54. Perez, D.L., Edwards, M.J., Nielsen, G., Kozlowska, K., Hallett, M., LaFrance, W.C., 2021a. Decade of progress in motor functional neurological disorder: continuing the momentum. J. Neurol. Neurosurg. Psychiatry jnnp-2020-323953. 10.1136/jnnp-2020-323953

55. Perez, D.L., Matin, N., Barsky, A., Costumero-Ramos, V., Makaretz, S.J., Young, S.S., Sepulcre, J., LaFrance, W.C., Keshavan, M.S., Dickerson, B.C., 2017. Cingulo-insular structural alterations associated with psychogenic symptoms, childhood abuse and PTSD in functional neurological disorders. J. Neurol. Neurosurg. Psychiatry 88, 491–497. 10.1136/jnnp-2016-314998

56. Perez, D.L., Nicholson, T.R., Asadi-Pooya, A.A., Bègue, I., Butler, M., Carson, A.J., David, A.S., Deeley, Q., Diez, I., Edwards, M.J., Espay, A.J., Gelauff, J.M., Hallett, M., Horovitz, S.G., Jungilligens, J., Kanaan, R.A.A., Tijssen, M.A.J., Kozlowska, K., LaFaver, K., LaFrance, W.C., Lidstone, S.C., Marapin, R.S., Maurer, C.W., Modirrousta, M., Reinders, A.A.T.S., Sojka, P., Staab, J.P., Stone, J., Szaflarski, J.P., Aybek, S., 2021b. Neuroimaging in Functional Neurological Disorder: State of the Field and Research Agenda. NeuroImage Clin. 30, 102623. 10.1016/j.nicl.2021.102623

57. Pick, S., Rojas-Aguiluz, M., Butler, M., Mulrenan, H., Nicholson, T.R., Goldstein, L.H., 2020. Dissociation and interoception in functional neurological disorder. Cognit. Neuropsychiatry 25, 294–311. 10.1080/13546805.2020.1791061

58. Ponnusamy, A., Marques, J.L.B., Reuber, M., 2012. Comparison of heart rate variability parameters during complex partial seizures and psychogenic nonepileptic seizures. Epilepsia 53, 1314–1321. 10.1111/j.1528-1167.2012.03518.x

59. Ponnusamy, A., Marques, J.L.B., Reuber, M., 2011. Heart rate variability measures as biomarkers in patients with psychogenic nonepileptic seizures: potential and limitations. Epilepsy Behav. EB 22, 685–691. 10.1016/j.yebeh.2011.08.020

60. Qian, J., Diez, I., Ortiz-Terán, L., Bonadio, C., Liddell, T., Goñi, J., Sepulcre, J., 2018. Positive Connectivity Predicts the Dynamic Intrinsic Topology of the Human Brain Network. Front. Syst. Neurosci. 12, 38. 10.3389/fnsys.2018.00038

61. Quadt, L., Critchley, H., Nagai, Y., 2022. Cognition, emotion, and the central autonomic network. Auton. Neurosci. Basic Clin. 238, 102948. 10.1016/j.autneu.2022.102948

62. Quigley, K.S., Berntson, G.G., 1996. Autonomic interactions and chronotropic control of the heart: heart period versus heart rate. Psychophysiology 33, 605–611. 10.1111/j.1469-8986.1996.tb02438.x

63. Radmanesh, M., Jalili, M., Kozlowska, K., 2020. Activation of Functional Brain Networks in Children With Psychogenic Non-epileptic Seizures. Front. Hum. Neurosci. 14, 339. 10.3389/fnhum.2020.00339

64. Rai, S., Foster, S., Griffiths, K.R., Breukelaar, I.A., Kozlowska, K., Korgaonkar, M.S., 2022. Altered resting-state neural networks in children and adolescents with functional neurological disorder. NeuroImage Clin. 35, 103110. 10.1016/j.nicl.2022.103110

65. Ricciardi, L., Demartini, B., Crucianelli, L., Edwards, M.J., Fotopoulou, A., 2014. INTEROCEPTIVE SENSITIVITY AND SENSE OF BODY OWNERSHIP IN PATIENTS WITH FUNCTIONAL NEUROLOGICAL SYMPTOMS. J. Neurol. Neurosurg. Psychiatry 85, e3–e3. 10.1136/jnnp-2014-308883.39

66. Rief, W., Hiller, W., 2003. A new approach to the assessment of the treatment effects of somatoform disorders. Psychosomatics 44, 492–498. 10.1176/appi.psy.44.6.492

67. Roberts, N.A., Burleson, M.H., Torres, D.L., Parkhurst, D.K., Garrett, R., Mitchell, L.B., Duncan, C.J., Mintert, M., Wang, N.C., 2020. Emotional Reactivity as a Vulnerability for Psychogenic Nonepileptic Seizures? Responses While Reliving Specific Emotions. J. Neuropsychiatry Clin. Neurosci. 32, 95–100. 10.1176/appi.neuropsych.19040084

68. Roberts, N.A., Burleson, M.H., Weber, D.J., Larson, A., Sergeant, K., Devine, M.J., Vincelette, T.M., Wang, N.C., 2012. Emotion in psychogenic nonepileptic seizures: responses to affective pictures. Epilepsy Behav. EB 24, 107–115. 10.1016/j.yebeh.2012.03.018

69. Saper, C.B., 2002. The central autonomic nervous system: conscious visceral perception and autonomic pattern generation. Annu. Rev. Neurosci. 25, 433–469. 10.1146/annurev.neuro.25.032502.111311

70. Sawchuk, T., Buchhalter, J., Senft, B., 2020. Psychogenic non-epileptic seizures in children - psychophysiology & dissociative characteristics. Psychiatry Res. 294, 113544. 10.1016/j.psychres.2020.113544

71. Schneider, A., Weber, S., Wyss, A., Loukas, S., Aybek, S., 2024. BOLD signal variability as potential new biomarker of functional neurological disorders. NeuroImage Clin. 43, 103625. 10.1016/j.nicl.2024.103625

72. Seeley, W.W., 2019. The Salience Network: A Neural System for Perceiving and Responding to Homeostatic Demands. J. Neurosci. Off. J. Soc. Neurosci. 39, 9878–9882. 10.1523/JNEUROSCI.1138-17.2019

73. Sepulcre, J., Sabuncu, M.R., Yeo, T.B., Liu, H., Johnson, K.A., 2012. Stepwise connectivity of the modal cortex reveals the multimodal organization of the human brain. J. Neurosci. Off. J. Soc. Neurosci. 32, 10649–10661. 10.1523/JNEUROSCI.0759-12.2012

74. Sojka, P., Diez, I., Bareš, M., Perez, D.L., 2021. Individual differences in interoceptive accuracy and prediction error in motor functional neurological disorders: A DTI study. Hum. Brain Mapp. 42, 1434–1445. 10.1002/hbm.25304

75. Sojka, P., Serranová, T., Khalsa, S.S., Perez, D.L., Diez, I., 2025. Altered Neural Processing of Interoception in Patients With Functional Neurological Disorder: A Task-Based fMRI Study. J. Neuropsychiatry Clin. Neurosci. 37, 149–159. 10.1176/appi.neuropsych.20240070

76. Spielberger, C.D., 2010. State-Trait Anxiety Inventory, in: Weiner, I.B., Craighead, W.E. (Eds.), The Corsini Encyclopedia of Psychology. Wiley, pp. 1–1. 10.1002/9780470479216.corpsy0943

77. Stephen, C.D., Fung, V., Perez, D.L., Espay, A.J., 2025. Comparison of Inpatient and Emergency Department Costs to Research Funding for Functional Neurologic Disorder: An Economic Analysis. Neurology 104, e213445. 10.1212/WNL.0000000000213445

78. Stevens, S.K., Williams, D.P., Thayer, J.F., Zalta, A.K., 2023. Differential Associations of Childhood Abuse and Neglect With Adult Autonomic Regulation and Mood-Related Pathology. Psychosom. Med. 85, 682–690. 10.1097/PSY.0000000000001239

79. Stürmer, M., Klinger-König, J., Vollmer, M., Weihs, A., Frenzel, S., Dörr, M., Kaderali, L., Felix, S.B., Stubbe, B., Ewert, R., Völzke, H., Grabe, H.J., Krause, E., 2025. Long-term alteration of heart rate variability following childhood maltreatment: Results of a general population study. Eur. Psychiatry J. Assoc. Eur. Psychiatr. 68, e86. 10.1192/j.eurpsy.2025.10040

80. Tarvainen, M.P., Niskanen, J.-P., Lipponen, J.A., Ranta-Aho, P.O., Karjalainen, P.A., 2014. Kubios HRV--heart rate variability analysis software. Comput. Methods Programs Biomed. 113, 210–220. 10.1016/j.cmpb.2013.07.024

81. Tinazzi, M., Morgante, F., Marcuzzo, E., Erro, R., Barone, P., Ceravolo, R., Mazzucchi, S., Pilotto, A., Padovani, A., Romito, L.M., Eleopra, R., Zappia, M., Nicoletti, A., Dallocchio, C., Arbasino, C., Bono, F., Pascarella, A., Demartini, B., Gambini, O., Modugno, N., Olivola, E., Di Stefano, V., Albanese, A., Ferrazzano, G., Tessitore, A., Zibetti, M., Calandra-Buonaura, G., Petracca, M., Esposito, M., Pisani, A., Manganotti, P., Stocchi, F., Coletti Moja, M., Antonini, A., Defazio, G., Geroin, C., 2020. Clinical Correlates of Functional Motor Disorders: An Italian Multicenter Study. Mov. Disord. Clin. Pract. 7, 920–929. 10.1002/mdc3.13077

82. Valenza, G., Sclocco, R., Duggento, A., Passamonti, L., Napadow, V., Barbieri, R., Toschi, N., 2019. The central autonomic network at rest: Uncovering functional MRI correlates of time-varying autonomic outflow. NeuroImage 197, 383–390. 10.1016/j.neuroimage.2019.04.075

83. van der Kruijs, S.J.M., Bodde, N.M.G., Vaessen, M.J., Lazeron, R.H.C., Vonck, K., Boon, P., Hofman, P.A.M., Backes, W.H., Aldenkamp, A.P., Jansen, J.F.A., 2012. Functional connectivity of dissociation in patients with psychogenic non-epileptic seizures. J. Neurol. Neurosurg. Psychiatry 83, 239–247. 10.1136/jnnp-2011-300776

84. Voon, V., Brezing, C., Gallea, C., Hallett, M., 2011. Aberrant supplementary motor complex and limbic activity during motor preparation in motor conversion disorder. Mov. Disord. Off. J. Mov. Disord. Soc. 26, 2396–2403. 10.1002/mds.23890

85. Walpola, I.C., Mohan, A., Foster, S., Kozlowska, K., 2025. Altered self-processing brain networks in paediatric functional neurological disorder. NeuroImage Clin. 47, 103811. 10.1016/j.nicl.2025.103811

86. Weber, S., Heim, S., Richiardi, J., Van De Ville, D., Serranová, T., Jech, R., Marapin, R.S., Tijssen, M.A.J., Aybek, S., 2022. Multi-centre classification of functional neurological disorders based on resting-state functional connectivity. NeuroImage Clin. 35, 103090. 10.1016/j.nicl.2022.103090

87. Wegrzyk, J., Kebets, V., Richiardi, J., Galli, S., de Ville, D.V., Aybek, S., 2018. Identifying motor functional neurological disorder using resting-state functional connectivity. NeuroImage Clin. 17, 163–168. 10.1016/j.nicl.2017.10.012

88. Westlin, C., Guthrie, A.J., Bleier, C., Finkelstein, S.A., Maggio, J., Ranford, J., MacLean, J., Godena, E., Millstein, D., Paredes-Echeverri, S., Freeburn, J., Adams, C., Stephen, C.D., Diez, I., Perez, D.L., 2025a. Delineating network integration and segregation in the pathophysiology of functional neurological disorder. Brain Commun. 7, fcaf195. 10.1093/braincomms/fcaf195

89. Westlin, C., Guthrie, A.J., Bleier, C., Finkelstein, S.A., Maggio, J., Ranford, J., MacLean, J., Godena, E., Millstein, D., Freeburn, J., Adams, C., Stephen, C.D., Diez, I., Perez, D.L., Katsumi, Y., 2025b. Functional Connectivity Gradients Reveal Altered Hierarchical Cortical Organization in Functional Neurological Disorder. Biol. Psychiatry Cogn. Neurosci. Neuroimaging S2451-9022(25)00325–8. 10.1016/j.bpsc.2025.10.010

90. Westlin, C., Guthrie, A.J., Paredes-Echeverri, S., Maggio, J., Finkelstein, S., Godena, E., Millstein, D., MacLean, J., Ranford, J., Freeburn, J., Adams, C., Stephen, C., Diez, I., Perez, D.L., 2025c. Machine learning classification of functional neurological disorder using structural brain MRI features. J. Neurol. Neurosurg. Psychiatry 96, 249–257. 10.1136/jnnp-2024-333499

91. Yeo, B.T.T., Krienen, F.M., Sepulcre, J., Sabuncu, M.R., Lashkari, D., Hollinshead, M., Roffman, J.L., Smoller, J.W., Zöllei, L., Polimeni, J.R., Fischl, B., Liu, H., Buckner, R.L., 2011. The organization of the human cerebral cortex estimated by intrinsic functional connectivity. J. Neurophysiol. 106, 1125–1165. 10.1152/jn.00338.2011

92. Zhang, J., Chen, D., Deming, P., Srirangarajan, T., Theriault, J., Kragel, P.A., Hartley, L., Lee, K.M., McVeigh, K., Wager, T.D., Wald, L.L., Satpute, A.B., Quigley, K.S., Whitfield-Gabrieli, S., Barrett, L.F., Bianciardi, M., 2025. Cortical and subcortical mapping of the allostatic-interoceptive system in the human brain using 7 Tesla fMRI. BioRxiv Prepr. Serv. Biol. 2023.07.20.548178. 10.1101/2023.07.20.548178

