## Supplementary Table 1 for "Relationships Between Brain Functional Connectivity and Resting Cardiac Autonomic Profiles in Functional Neurological Disorder: A Pilot Study"

**SUPPLEMENTARY TABLES**

**Supplementary Table 1. Demographic characteristics of participants with functional neurological disorder (FND) and psychiatric controls.**

| **Subject** | **Phenotype** | **Phenotypic Description** | **Current SCID-DSM-5 Diagnoses** | **Past SCID-DSM-5 Diagnoses** | **Psychotropic Medications** |
| --- | --- | --- | --- | --- | --- |
| 1 | FND-Motor | clinically-established functional tremor, functional dystonia & functional gait | PD+AG, SSD | GAD, DEP NOS | - |
| 2 | FND-Motor | clinically-established functional facial spasms/tics & functional speech | GAD, PD+AG, ADHD, PTSD | MDE | CLN, LDA, APR, SERT, LRZ |
| 3 | FND-Motor | clinically-established functional gait & functional speech | - | DEP NOS, ANX NOS | GBP |
| 4 | FND-Motor | clinically-established functional jerks/spasms/tics | PTSD, ADHD | SAD, MDE, Specific Phobia | MIR, SERT, MPD, LRZ |
| 5 | FND-Seizure | documented functional seizures | GAD, ADHD | MDE, AG | LTG, NRT, FLX, MPD |
| 6 | FND-Motor | clinically-established functional limb weakness (right arm/leg) & functional speech | PTSD, GAD, MDE, PD+AG, SSD | MDE | APR, FLX, GBP, LRZ, MLT |
| 7 | FND-Motor | clinically-established functional tremor & functional dystonia (right foot) | MDE, GAD, PTSD | MDE, PD+AG, SAD | BSP |
| 8 | FND-Seizure | documented functional seizures | GAD, PTSD, ADHD | Eating Disorder, MDE, PD+AG | OLZ, CTP, APM / DXAM |
| 9 | FND-Seizure | documented functional seizures | MDE, GAD, SAD | Eating Disorder | MIR, CLP |
| 10 | FND-Seizure, FND-Motor | probable functional seizures; clinically-established functional limb weakness (bilateral leg), functional tremor, & functional gait | SSD, AG, DYS | MDE, PD-AG, GAD, PTSD, Eating Disorder | AMT, DLX |
| 11 | FND-Motor | clinically-established functional dystonia & functional speech | GAD | PTSD | - |
| 12 | FND-Motor | clinically-established functional tremor | PTSD, MDE, Anxiety NOS | - | MIR, CLP |
| 13 | FND-Motor | clinically established functional tremor | PTSD, MDE, SAD, GAD, SSD | MDE, PTSD, SUD, Anxiety NOS | APR, BUS, PZN, ECP, TZD, HDZ |
| 14 | FND-Seizure, FND-Motor | documented functional seizures; clinically-established functional jerks/tics, & functional speech | PD+AG, DYS, GAD, SSD | OCD | DLX, GBP, LRZ |
| 15 | FND-Seizure | documented functional seizures | AG, SAD, OCD, SSD | - | HDZ, DZP |
| 16 | FND-Seizure | documented functional seizures | PTSD, GAD, SSD, ADHD | MDE | AMP / DXAM, PZN, CTP |
| 17 | FND-Motor | clinically established functional gait & functional speech | GAD | MDE | CTP, CLN, TZD |
| 18 | FND-Motor | clinically established functional limb weakness, functional tremor, functional jerks and functional gait | SSD | - | - |
| 19 | FND-Motor | clinically established functional gait, functional limb weakness (left-leg), functional tremor, functional speech | Specific Phobia | MDE | - |
| 20 | FND-Motor | clinically-established functional limb weakness (left arm and leg) & functional gait | - | MDE, SAD, PTSD, Eating Disorder | CTP |
| 21 | Psychiatric Control | - | - | PTSD, MDE | - |
| 22 | Psychiatric Control | - | - | DEP NOS | - |
| 23 | Psychiatric Control | - | GAD | MDE | - |
| 24 | Psychiatric Control | - | - | MDE | GBP, CTP |
| 25 | Psychiatric Control | - | Specific Phobia | - | LRZ |
| 26 | Psychiatric Control | - | PTSD, GAD, MDE, DYS | Eating Disorder | VNF, MPD, LRZ, PZN, QTP, GBP, BPN, EZP |
| 27 | Psychiatric Control | - | - | OCD, GAD | ECP |
| 28 | Psychiatric Control | - | PTSD, OCD, SAD | MDE | FLX |
| 29 | Psychiatric Control | - | ANX NOS* | - | LDA |
| 30 | Psychiatric Control | - | - | MDE, GAD | - |
| 31 | Psychiatric Control | - | ANX NOS, SSD | MDE, SAD | BUP |
| 32 | Psychiatric Control | - | PTSD, MDE, GAD | - | SERT, ECP, TZD, APZ |
| 33 | Psychiatric Control | - | MDE, GAD, PTSD, ADHD | MDE, Eating Disorder | LDA, LTG, LRZ, CLN |
| 34 | Psychiatric Control | - | - | MDE | BUP, APM / DXAM, TZD, VNF |
| 35 | Psychiatric Control | - | MDE, SAD | PTSD, SUD | CTP, MPH |
| 36 | Psychiatric Control | - | - | MDE, PTSD | - |
| 37 | Psychiatric Control | - | PTSD, MDE | - | - |
| 38 | Psychiatric Control | - | SAD, PD + AG, MDE | - | VNF, SERT, HDZ |
| 39 | Psychiatric Control | - | PD, OCD | Eating Disorder, MDE, SAD | LDA, ECP |
| 40 | Psychiatric Control | - | - | GAD | FLX, LDA |
| 41 | Psychiatric Control | - | GAD | MDE | - |
| 42 | Psychiatric Control | - | GAD, Specific Phobia, ADHD | MDE | ATX |
| 43 | Psychiatric Control | - | DEP NOS, ANX NOS, Specific Phobia | MDE, AG | VNF, BUP, LRZ, APM / DXAM |

FND-Motor, Functional Motor Disorder; FND-Seiz, Functional Seizures; SCID-DSM-5, Structured Clinical Interview for DSM 5^th^ Edition [note subject 1 received a SCID-I for DSM-IV]; ADHD, Attention-Deficit/Hyperactivity Disorder; AG, Agoraphobia; ANX, Anxiety; AUD, Alcohol Use Disorder; BPAD, Bipolar Affective Disorder; DEP, Depression; DYS, Dysthymia; GAD, Generalized Anxiety Disorder; IAD, Illness Anxiety Disorder; MDE, Major Depressive Episode; NOS, not otherwise specified; OCD, Obsessive Compulsive Disorder; PD+AG, Panic Disorder with Agoraphobia; PD-AG, Panic Disorder without Agoraphobia; PTSD, Post-Traumatic Stress Disorder; SAD, Social Anxiety Disorder; SSD, Somatic Symptom Disorder; SUD, Substance Use Disorder; AMA, Amantadine; AMT, Amitriptyline; APM, Amphetamine; APR, Aripiprazole; APZ, Alprazolam; ATX, Atomoxetine; BUP, Bupropion; BUS, Buspirone; CBD, Cannabidiol; CLN, Clonidine; CLP, Clonazepam; CTP, Citalopram; DLX, Duloxetine; DXAM, Dextroamphetamine; DZP, Diazepam; ECP, Escitalopram; FLX, Fluoxetine; GBP, Gabapentin; HDZ, Hydroxyzine; LDA, Lisdexamfetamine; LTM, Lithium; LTG, Lamotrigine; LRZ, Lorazepam; MIR, Mirtazapine; MPH, Methylphenidate; NRT, Nortriptyline; PGB, Pregabalin; PZN, Prazosin; QTP, Quetiapine; SERT, Sertraline; TPM, Topiramate; TZD, Trazodone; VNF, Venlafaxine.

*SCID data were unavailable; psychiatric diagnosis was based on screening measures.
