## Supplementary Table 2 for "Relationships Between Brain Functional Connectivity and Resting Cardiac Autonomic Profiles in Functional Neurological Disorder: A Pilot Study"

**Supplementary Table 2. Resting Photoplethysmography (PPG) Segment-Level Quality and Signal Characteristics from Empatica E4 Recording.**

|  | **Functional Neurological Disorder**  **(n=20)**  mean ± SD | **Psychiatric  Controls**  **(n=23)**  mean ± SD | **p-value** ^a^ |
| --- | --- | --- | --- |
| Number of PPG segments analyzed per subject | 8.1 ± 1.7 | 8.1 ± 1.2 | 0.71 (0.71) |
| Segment duration (s) | 58.4 ± 4.6 | 58.0 ± 4.5 | 0.69 (0.71) |
| Total analyzed time (s) | 473.1 ± 110.9 | 472.0 ± 78.6 | 0.58 (0.71) |
| Respiration rate (Hz) | 0.2 ± 0.1 | 0.3 ± 0.1 | 0.64 (0.71) |
| Segments with respiration rate outside HF band (%) | 7.5 ± 20.5 | 11.2 ± 23.4 | 0.31 (0.71) |

Each “segment” corresponds to a 40–60 s resting PPG window extracted from Empatica E4 recordings and processed using *Kubios HRV Scientific* (version 4.1.2.1). HF band defined as 0.15-0.40 Hz. ^a^ *p*-values correspond to uncorrected statistics, with false discovery rate (FDR)-corrected *p*-values shown in parentheses.
