## Supplementary figures and images for "Relationships Between Brain Functional Connectivity and Resting Cardiac Autonomic Profiles in Functional Neurological Disorder: A Pilot Study"

### Supplementary Figure 1

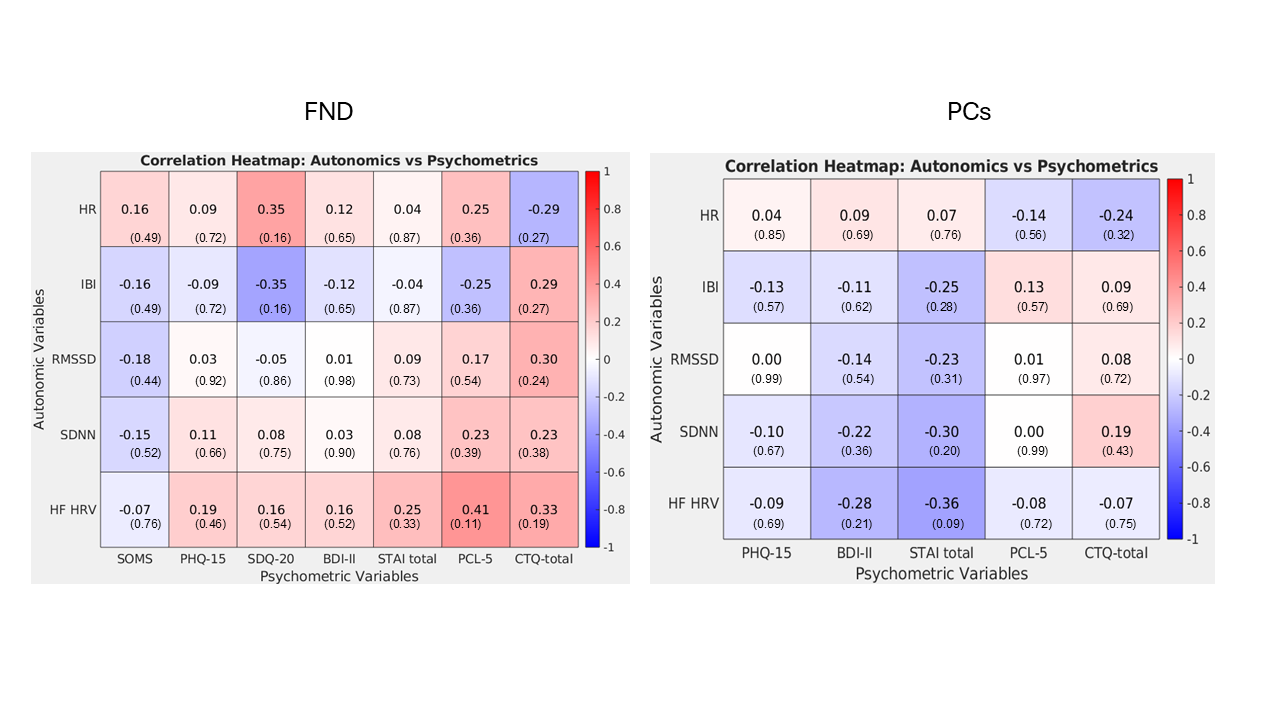

### Supplementary Figure 2

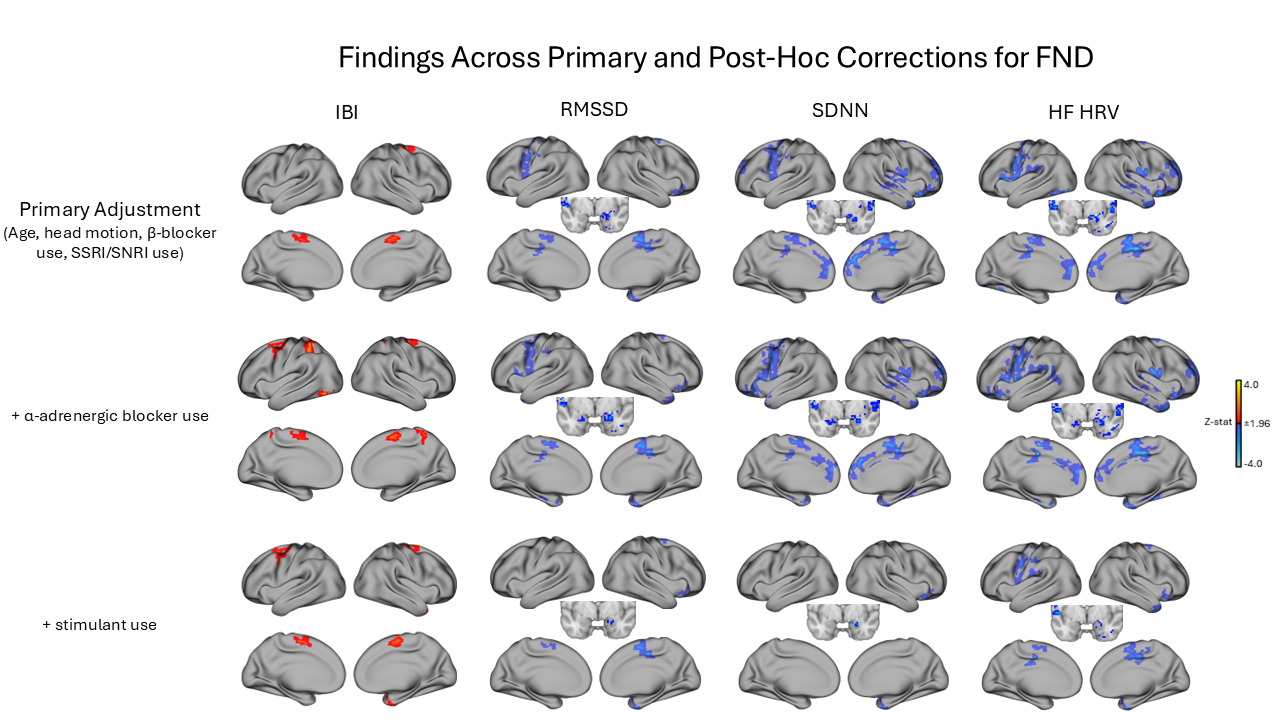

### Supplementary Figure 3

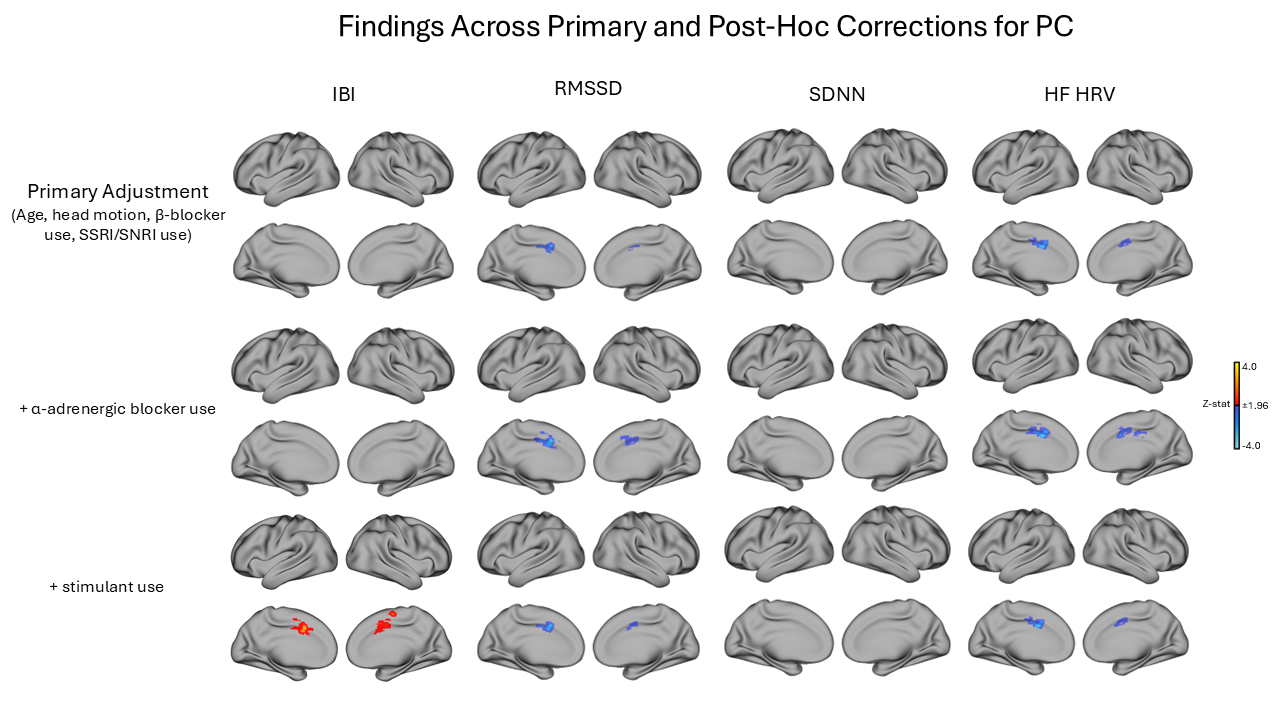
